# A phase 1 randomized controlled trial to evaluate the safety and immunogenicity of a HIV monomeric gp120 protein B-cell lineage targeting HIV vaccine in healthy adults

**DOI:** 10.64898/2026.05.26.26353896

**Authors:** James J. Kobie, Wilton B. Williams, William O. Hahn, Paul T. Edlefsen, Margaret Brewinski Isaacs, Maurine D. Miner, K. Rachael Parks, Stephen C De Rosa, Huijun An, Claudio Yurdadon, Jordan Spreng, Jongln Hwang, Matthew Clark, Vaibhav Jain, Simon G. Gregory, Madison Berry, Kevin Wiehe, Paul A. Goepfert, Hong-Van Tieu, Michael C. Keefer, Lindsey R. Baden, Spyros Kalams, Cecilia Morgan, David C. Montefiori, Guido Ferrari, Stephanie Regenold, Georgia D. Tomaras, M. Juliana McElrath, Lawrence Corey, Magdalena E. Sobieszczyk, Barton F. Haynes, HVTN 115 Study Team

**Affiliations:** University of Alabama at Birmingham, Birmingham, AL; Duke Human Vaccine Institute, Duke University School of Medicine, Durham, NC; Vaccine and Infectious Disease Division, Fred Hutchinson Cancer Center, Seattle, WA; Division of AIDS, National Institute of Allergy and Infectious Diseases, Bethesda, MD, USA; The Duke Molecular Physiology Institute, Duke University, Durham, NC; Laboratory of Infectious Disease Prevention, Lindsley F. Kimball Research Institute, New York Blood Center, New York, NY, Columbia University Irving Medical Center, New York, NY; Department of Medicine, University of Rochester, Rochester, NY; Division of Infectious Diseases, Brigham & Women’s Hospital, Boston, MA; Vanderbilt University, Nashville, TN; Duke University School of Medicine, Durham, NC; Division of Infectious Diseases, Columbia University Irving Medical Center, Aaron Diamond AIDS Research Center, Vagelos College of Physicians and Surgeons, New York, NY

**Author notes:** co-first authors (James J. Kobie, Wilton B. Williams). co-senior authors (Magdalena E. Sobieszczyk, Barton F. Haynes). Corresponding author; James Kobie, University of Alabama, Birmingham.

## Abstract

**Background:** The isolation of many HIV broadly neutralizing antibodies (bnAbs) from people living with HIV (PLWH) and rigorous characterization of their ontogeny has promoted the goal of reverse engineering their natural development as a strategy for achieving an effective preventive HIV vaccine. We previously described the developmental process of CH103, a CD4-binding site (CD4bs)-specific monoclonal antibody, and the associated evolution of HIV Envelopes (Envs) within the person (CH505) from whom it was isolated. A series of monomeric gp120 protein subunit immunogens representing the transmitted founder (TF) and Envs that evolved during infection and optimally reacted with lineage members at each step of the CH103 clone maturation path were evaluated in this placebo controlled randomized vaccine trial to test--for the first time in humans--the concept of whether sequential immunization with gp120 monomeric proteins can recapitulate the development of CD4bs B-cell clonal lineages, including CH103.

**Methods:** HIV Vaccine Trials Network 115 (HVTN 115) was a randomized placebo-controlled vaccine trial at US clinical research sites. We tested the safety and immunogenicity of CH505TF gp120 + GLA-SE (Part A), and then the ability of sequential CH505 gp120 proteins (corresponding to CH505’s weeks 53 and 78 Envs) + GLA-SE immunizations to induce CD4bs-specific neutralizing antibodies (Part B). We assessed binding and neutralizing antibody responses, antibody dependent cellular cytotoxicity, antibody dependent cellular phagocytosis, T-cell responses and B-cell phenotyping.

**Results:** We enrolled 42 participants between October 2017 and May 2018 for Part A, and 65 participants from December 2020 to October 2022 for Part B. Immunization with the CH505 gp120 proteins adjuvanted with GLA-SE was well tolerated and induced CD4bs-specific B cells and Env-specific plasma antibodies. The plasma neutralizing antibody response was limited to primarily tier 1 autologous and heterologous HIV-1 strains. Blood-derived B-cell repertoire analyses identified CD4bs antibodies that preferentially bound to open-occluded trimeric Envs that exist in an intermediate state between prefusion-closed to CD4-bound open confirmations, consistent with tier 1 HIV neutralizing activity.

**Conclusions:** Together, these results suggest that the low-affinity CH505TF gp120 monomer elicited CD4bs antibodies in the sera and B-cell repertoires of humans. However, our findings also indicate that gp120 monomers are insufficient to induce detectable bnAb precursors to epitopes on native Env trimers. Nonetheless, our data provide a benchmark for comparison with ongoing clinical trials testing high-affinity CH505 Env trimers for induction of CD4bs bnAb precursors.

## INTRODUCTION

Despite the high global burden of disease and tremendous research efforts, an HIV vaccine remains elusive. Monoclonal antibodies can prevent acquisition of HIV against susceptible strains ^1^. An ideal HIV vaccine candidate would elicit a neutralizing antibody response effective against numerous strains of HIV (i.e., “broadly neutralizing” or bnAbs). BnAbs have been observed repeatedly in people living with HIV (PLWH) for long time periods ^2^. Unfortunately, development of these bnAbs during chronic HIV can take years, as it typically requires extensive B cell somatic mutation and evolution^3^. Vaccines developed thus far have failed to elicit serum with antibodies that have the necessary potency and breadth to protect against HIV acquisition.

One hypothesis for the absence of bnAbs observed following repeated HIV vaccination with the same Env is that the virus changes in response to antibody-mediated immune pressure during chronic infection. As a result, the HIV envelopes (Envs) present during late or chronic infections may be very different than the Env of early viruses due to pressure from the immune system. Additionally, previous studies in PLWH have implied that it is a variant of the transmitted-founder (TF) Env that bound the unmutated common ancestor (UCA) of the B cell lineages that ultimately produce mature bnAbs^4^. Thus, careful investigations into the evolution of host and B cell responses shortly after infection have illustrated that Env variants present early during infection guide B cell maturation in a way that ultimately results in bnAb-producing B cells^4,5^.

The current study was designed to evaluate whether it is possible to stimulate the germline or UCA of a bnAb lineage using vaccination, guiding the evolutionary development of B cells using sequential Env immunogens that recapitulate the natural evolution of the HIV Env in PLWH. The concept is that a vaccination series would start with the TF strain and then progress using Env variants that were isolated during chronic infection from an individual who developed bnAbs, with the goal of recreating the sequence of events that lead to the presence of broadly neutralizing antibodies in serum.

To initiate bnAb lineages, the recombinant Env must contain epitopes that bind to the UCA (naïve B cell receptors) of bnAb lineages^6,7^. The bnAb CH103, targeting the Env CD4 binding site (CD4bs), was isolated from a PLWH (identification number CH505) for whom both plasma and viral samples were available from the time of acquisition and for several years following initial acquisition^5^. It was therefore possible to sequentially map the TF Env and cognate B cell throughout B cell maturation and intra-host viral coevolution. A single TF virus was able to bind to the CH103 UCA, and a series of evolved envelope proteins of the founder virus that were likely stimulators of the bnAb lineage were identified^5^.

Gao and colleagues identified in the same CH505 individual an additional CD4-binding site bnAb lineage termed CH235. CH505 M5 is a natural mutant of CH505 TF with a single N279K change that occurred very early after infection in the CH505 individual^8^. Initially, Gao and colleagues found that CH505M5 gp120 bound recombinant mAbs representative of the earliest members of the CH235 lineage, suggesting that CH505 M5 is a promising immunogen for inducing the CH235 bnAb lineage^9,10^.

At the time of the initial design of this trial, it was unknown whether an Env monomer gp120 subunit would be sufficient to prime naïve B cells in humans. Therefore, a series of monomeric gp120 protein subunit immunogens representing the TF Env (CH505 TF), and that optimally reacted with each step of the CH103 lineage from weeks 53, 78, and 100 following acquisition, were developed to evaluate the concept of whether sequential immunizations are able to recapitulate the development of the CH103 lineage. Sequential immunization of rhesus macaques with CH505 gp120 Envs resulted in improved neutralizing antibody breadth, albeit for easy-to-neutralize, lab-adapted tier 1 HIV strains, as compared to repeated immunization with CH505 TF gp120 alone^8^.

Herein, we present the results of HVTN 115 as a first in human clinical trial for testing the concept of whether sequential HIV Env immunizations can recreate B-cell evolution associated with induction of CD4bs neutralizing antibody lineages, including CH103 bnAb precursors. The trial was designed to first determine the safety and immunogenicity of different doses of CH505TF gp120 (Part A), and then the ability of sequential CH505 gp120 protein immunizations to induce CD4bs-specific neutralizing antibodies (Part B).

## MATERIALS AND METHODS

### Study design

HVTN 115 was a randomized, double-blind, placebo-controlled Phase 1 study conducted at four clinical research sites in the USA: University of Alabama at Birmingham (Birmingham, AL); University of Rochester (Rochester, NY); New York Blood Center (New York, NY); and Columbia University Physicians & Surgeons (New York, NY). Healthy adults living without HIV between the ages of 18 and 50 were screened for eligibility, including low likelihood of HIV acquisition. Participants were randomized 2:2:2:1 to receive CH505TF at 20 mcg (n=12, T1), 100 mcg (n=12, T2) or 400 mcg (n=12, T3), or placebo (n=6, T4) at months 0, 2, 4, 8 and 12 in Part A (**Figure 1**). In Part B, participants in T5 (n=20) received 400 mcg CH505TF at month 0, CD505w53 at month 2, and CH505w78 at months 4 and 8; in T6 (n=20) received 400 mcg CH505TF at month 0, CH505TF + CH505w53 at month 2, CH505TF + CH505w53 + CH505w78 at month 4, and CH505w53 + CH505w78 at month 8; in T7 (n=20) received 400 mcg CH505 M5 at months 0, 2, 4, and 8; and in T8 (n=5) received placebo at months 0, 2, 4, and 8 (**Figure 1)**.

**Figure 1.**
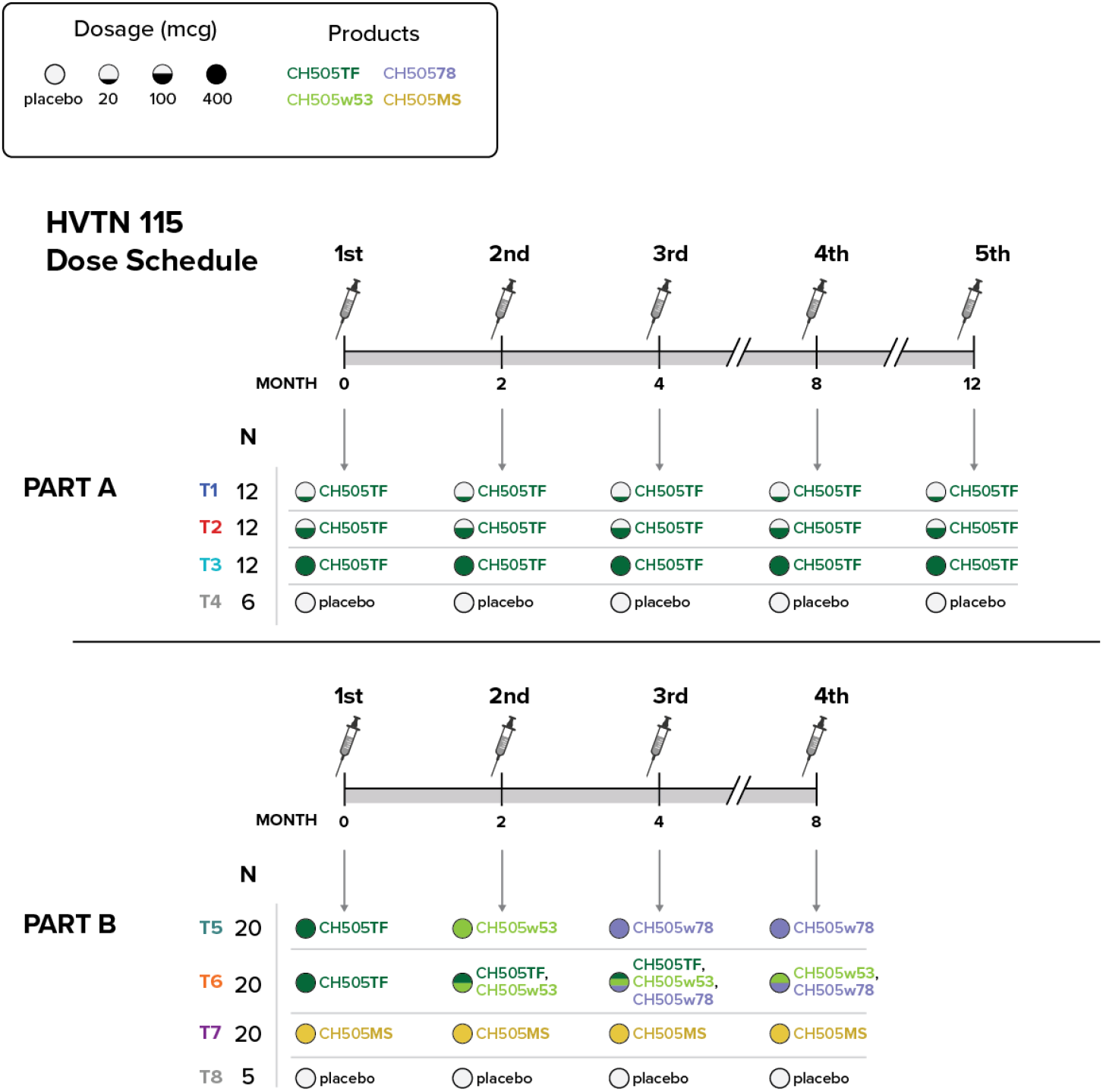
Schematic of study design.

The vaccine was a CH505TF gp120 transmitted founder HIV Env protein (DAIDS, NIAID, Bethesda, MD) with GLA-SE (glucopyranosyl lipid adjuvant-stable emulsion [synthetic lipid A derivative mixed with squalene oil]) adjuvant (AAHI, formerly IDRI, Seattle, WA). The total dose of GLA-SE was 10 mcg at all timepoints. The total volume for protein plus adjuvant for injection was 1 mL, mixed 1:1 by volume. Placebo was Sodium Chloride for Injection, 0.9%. All injections were administered intramuscularly (IM) in the thigh. Participants were followed in the clinic for a total of 24 months. Blood draws occurred during screening, month 0 (first vaccination) and months 0.25, 0.5, 2.5, 4.5, 8.5, 12, 12.25, 12.5, 15 and 18. Health contacts continued for one year following final injection of study product. All participants provided written informed consent, and the protocol was approved by each clinical site’s Institutional Review Board and central IRB and registered at ClinicalTrials.gov (NCT03220724). Trial safety of participants was overseen by the HVTN 115 Protocol Safety Review Team and HVTN Safety Monitoring Board.

The sample size was based on planned enrollments of each parts, allowing for 10% missing immunogenicity data (35% in Part C for leukapheresis) and chosen to provide adequate safety-event detection and ensuring the study met prespecified safety-event detection goals and immunogenicity power requirements for the primary comparisons based on prior HVTN data. The randomization sequence was parallel and permuted blocked and obtained by computer-generated random numbers and provided to each site through the web-based randomization system. The trial was double-blinded such that participants and site staff, including investigators, were blinded to treatment assignment, while only site pharmacists, selected protocol/pharmacy and monitoring staff, SDMC staff, and SMB members had access to assignments as needed, with blinding maintained by restricting assignment information to pharmacy and other authorized personnel and prohibiting disclosure except through prespecified emergency unblinding procedures. Sample size has accounted for 10% of missing data.

### Binding antibody multiplex assay (BAMA)

Serum HIV-1-specific IgA and IgG responses against antigens listed in **Supplementary Table 1** were measured (IgA responses were measured in IgG depleted serum) on a Bio-Plex instrument (Bio-Rad) using a standardized custom HIV-1 Luminex assay ^11–15^. The readout was background-subtracted mean fluorescence intensity (MFI), where background referred to a plate level control (i.e., a blank well run on each plate). Standard positive and negative controls were included in each assay. The IgA positive controls were purified polyclonal IgG from PLWH and CH103 IgG. Additional positive controls included 7B2 IgA as detection controls. The IgG positive controls were PGT145 IgG and VRC01 IgG. The negative controls included HIV-1 seronegative human sera and blank beads.

### FcR BAMA

The frequency and magnitude of FcGRIIIa F158 and FcgRIIa H131 binding antibody responses were measured by BAMA from serum specimens obtained at month 4.5, month 8.5, and month 12.5. FcR BAMA utilizes Fc Receptor (FcR) proteins tetramerized with phycoerythrin (PE) as a detection reagent to determine antigen specific binding antibody interactions with FcRs. Microspheres were read on a Flexmap 3D instrument. The readout was background-subtracted MFI, where background referred to a plate level control. The positive controls for gp120 antigens were purified polyclonal IgG from PLWH. The positive controls for SOSIP binding were PGT145 and VRC01. The negative controls included HIV-1 seronegative human sera and blank beads.

### Intracellular cytokine staining (ICS)

Endpoints were the response rate and the magnitude of T-cell responses as measured by the ICS assay from PBMC specimens obtained at month 4.5 and month 12.5. Flow cytometry was used to examine HIV-1-specific CD4+ and CD8+ T-cell responses using a validated ICS assay. A 17-color staining panel was used (**Supplementary Table 2**). The global potential T-cell epitope peptide pools (PTEg) evaluated were Env-1-PTEg-SEQ and Env-2-PTEg-SEQ.

Previously cryopreserved PBMC specimens were stimulated ex vivo with the synthetic peptide pools. As a negative control, cells were not stimulated. As a positive control, cells were stimulated with a polyclonal stimulant, staphylococcal enterotoxin B and with peptides covering key T-cell epitopes in the cytomegalovirus pp65 protein. There were no replicates except for the negative control, which had two replicates.

### Neutralizing antibody (nAb) responses

The frequency and magnitude of nAb responses were measured by the HIV-1 nAb assay from serum specimens obtained at 2 weeks post 3rd vaccination (month 4.5), 2 weeks post 4th vaccination (month 8.5), and 2 weeks post 5th vaccination (month 12.5). nAbs against HIV-1 were measured as a function of reductions in Tat-regulated luciferase (Luc) reporter gene expression in TZM-bl cells. The assay measured neutralization titers against a panel of autologous and heterologous Env-pseudotyped viruses produced in either 293T cells or 293S/GnT1-cells. 1) Autologous, tier 1A neutralization phenotype: CH0505.w4.3 (293T); 2) Autologous CH0505 Env lineage, tier 2 neutralization phenotype: CH0505s, CH0505.w53.16, CH0505.w78.e33, CH0505.w100.B6, CH0505TF.M5 (all 293T); 3) CH235 CD4bs bnAb precursor detection: CH0505s.G458Y.N279K.2 (GnT1-); 4) CH235 CD4bs bnAb precursor confirmation: CH0505s.G458Y.N279K.N280D.6 (GnT1-); 5) CH103 CD4bs bnAb precursor detection: CH0505TF.gly4 (GnT1-); and 6) CH01 V2-glycan bnAb precursor detection: SIVcpzMT145.Q171K, Q23.17. CH0505TF.M5: M5 refers to a naturally occurring N279K substitution that arose early during infection.

### Antibody-dependent cellular phagocytosis (ADCP)

The frequency and magnitude of average phagocytosis score responses were measured in serum specimens obtained at baseline (month 0), month 4.5, month 8.5, and month 12.5 for Part A.

To assess the ability of polyclonal IgG to mediate phagocytosis, ADCP was measured using methods previously described^16^. Biotinylated antigen conjugated to neutravidin beads were incubated with patient serum or plasma, purified IgG or monoclonal antibodies. A monocyte cell line (THP-1) was added and incubated with the antibody/bead mixture. Cells were then analyzed for bead internalization by flow cytometry (bead positive versus bead negative detection). Samples were tested in duplicate at 1:50 dilution for the CH505TF_D7gp120.avi/293F antigen. A phagocytic score was determined based on the ratio of experimental sample to PBS control. Mean phagocytosis score was defined as: (% bead positive for participant x MFI bead positive for participant) / (% bead positive for PBS only control x MFI bead positive for PBS only control). All samples including controls were run in duplicate within the assay. The following acceptance criteria were employed for this assay: 1. <30% CV between individual replicates within assay for positive responders; 2. Positive, negative and controls containing no antibody. Lower and upper limits of quantification were not defined for the CH505TF_D7gp120.avi/293F antigen at 1:50 dilution.

### B cell phenotyping

The frequency of CH505TF Env gp120 and CD4 binding site-specific B cells was measured by the flow cytometry assay from PBMC specimens obtained at baseline (day of 1st vaccination; Month 0), two weeks after the 3rd vaccination (Month 4.5), and two weeks after the 5th vaccination (Month 12.5). The CD4bs and Env-specific B cells were quantified using biotinylated CH505TF gp120 protein probes (conjugated with streptavidin-Alexa647 and streptavidin-BV711) and the CD4bs-mutant (CH505TF D7 gp120 IΔ371 labeled with streptavidin-phycoerythrin). The vaccine-matched CH505 proteins were designed, biotinylated, and provided by Kevin Saunders, Tony Moody and Barton Haynes, Duke University^5,8^. Live total B cells were identified using doublet exclusion, lymphocyte scatter profile, viability dye, and the following lineage markers: negative for CD3, CD56 and CD14, and positive for CD19 and CD20. IgG+ B cells were further gated on IgD negative and IgG+. The reported frequencies are: % CH505+ of total B cells equals the frequency of double-positive CH505 gp120+ B cells out of total B cells; % CD4bs CH505+ of total B cells equals the frequency of double-positive CH505 gp120+ B cells that are negative for the CD4bs mutant (CH505Δ371) out of total B cells; % CH505+ of IgG+ B cells equals the frequency of double-positive CH505 gp120+ IgG+ B cells out of IgG+ B cells; and % CD4bs CH505+ of IgG+ B cells equals the frequency of double-positive CH505 gp120+ IgG+ B cells that are negative for the CD4bs mutant (CH505Δ371) out of IgG+ B cells. Participant PBMCs were incubated with fluorescently labeled recombinant BG505-SOSIP.664 Env proteins and other fluorescently labeled antibodies^17^. Live total B cells were defined based on viability dye staining, scatter profile, and doublet exclusion. The following lineage markers were used to identify B cells: negative for CD3, CD56, and CD14, and positive for both CD19 and CD20. IgG^+^ B cells were further gated as IgD™ and IgG^+^. The percent of BG505 SOSIP.664 gp140–specific IgG^+^ B cells, out of the total number of IgG^+^ B cells, was reported. A positive response determination was based on a participant-level statistical comparison of Env-specific B cells after vaccination vs. baseline using a Fisher’s exact test.

### Infected cell antibody binding assay (ICABA)

The magnitude of responses was measured by the ICABA assay from presence of antibodies binding to the surface of HIV-1 infected cells in the serum of participants using Infectious Molecular Clone (IMC) infected target cells obtained at baseline and 2 weeks post 4th vaccination (M8.5) for Part B.

CEM.CCR5.NKR cells infected with HIV-1 IMC CH505 and mock-infected CEM.CCR5.NKR cells were tested in a 96-well plate for the ICABA. Participant sera in addition to controls were incubated with the IMC infected cells, and the presence of bound antibodies were detected on the surface of the cells using flow cytometry. The data from ICABA are quantitative.

### Antibody dependent cellular cytotoxicity (ADCC)

ADCC-mediated antibody responses were measured using Luc and GTL assays from specimens in Part A groups 1 and 4 obtained at visit 2 (baseline) and visit 13 (2 weeks post fifth study product administration). The GTL ADCC assay measures percent granzyme B activity, defined as the percentage of antigen coated target cells positive for proteolytically active granzyme B out of the total viable target cell population. Endpoints are the response rate and magnitude of ADCC mediated antibody responses against Clade B CH505 gp120. The Luciferase ADCC assay tested the presence of antibody responses against IMC infected target cells by measuring percent reduction in RLUs, reported as percentage specific killing. Endpoints are the response rate and the magnitude of ADCC responses against HIV IMC CH505.

The qualified GranToxiLux ADCC (ADCCGTL) assay was performed as previously described^18^. Target cells were a clonal isolate of the CEM.NKRCCR5 CD4+ T cell line^19^ coated with a recombinant gp120 representing the HIV1 envelope of the subtype Clade C CH505. Effector cells were PBMCs obtained from a HIV seronegative donor with heterozygous FcγR3A at position 158 (158F/V).

Peripheral blood mononuclear cells (PBMCs) were obtained by leukapheresis to collect sufficient cells for completion of the study with a single donation, minimizing potential effector cell population variability effects on the study outcome. PBMCs were used at an effector cell to target cell ratio of 30:1. Serum samples were tested after fivefold serial dilutions starting at 1:50. Each plate has one standardized positive control in duplicate and one standardized negative control in duplicate. ADCC is quantified as net percent granzyme B activity, which is the percent of target cells positive for GTL (an indicator of granzyme B uptake) minus the percent of target cells positive for GTL when incubated with effector cells in the absence of a source of antibodies. Flow cytometry is used to quantify the frequency of granzyme B positive cells. ADCC was quantified as net percent granzyme B activity, i.e. the percent of target cells positive for GTL minus the percent of target cells positive for GTL when incubated with effector cells in the absence of any source of antibodies. A modified version of a previously published ADCC luciferase procedure^20^ was utilized. Briefly, CEM.NKRCCR5 cells^19^ were used as targets for ADCC luciferase assays after infection by the following HIV1 IMC: HIV CH0505s.LucR.T2A.ecto/293T/17 (accession number KC247557; abbreviated name HIV IMC CH505; vaccine matched).

PBMCs were obtained from an HIV seronegative donor by leukapheresis and cryopreserved until the day of the assay. After thawing and overnight resting in RPMI 1640 supplemented with antibiotics, 10% fetal bovine serum (R10), and 10 ng/mL of IL15, the PBMCs were used as effector cells at an effector to target ratio of 30:1.

Target and effector cells were plated in white 96-well half area plates and cocultured with 4-fold serial dilutions of trial participant serum starting at the 1:50 dilution. For each sample, percent specific killing was measured in duplicate at dilutions of 1:50, 1:200, 1:800. Cocultures were incubated for 6 hours at 37°C in 5% CO2. The final readout was the reduction of luminescence intensity generated by the presence of residual intact target cells that had not been lysed by the effector population in the presence of ADCC mediating serum antibodies. The percentage of killing was calculated using the formula: percent specific loss Luciferase activity = 100* (RLU of target and effector well– RLU of test well)/(RLU of target and effector well).

In this analysis, the RLU of the target plus effector wells represents spontaneous lysis in the absence of any source of antibody and is used to calculate background activity. The monoclonal antibody (Synagis) and a cocktail of HIV-1 monoclonal antibodies (A32, 2G12, CH44, and 7B2) were used as negative and positive controls, respectively.

### ELISA binding

Monoclonal antibody binding was measured by ELISA in 384-well ELISA plates (Costar #3700) coated with 30ng protein antigen in 0.1M sodium bicarbonate overnight at 4°C as previously described^21,22^. ELISA binding data were collected by Spectramax Plus384 plate reader, analyzed using Softmax Pro version 5.3, and collated or graphed using excel and GraphPad Prism software.

### PCR isolation of antibodies from antigen-specific B cell sorts

Single sorted B cells were obtained from HVTN 115 vaccine recipients using previously described flow cytometry-based sorts^8,23^. Cells were individually sorted into 96-well plates containing lysis buffer and immediately stored at −80°C. Human V_H_ DJ_H_ and V_L_ J_L_ segments were isolated by single-cell PCR^8,23^. Antibody sequences were analyzed using a custom-built bioinformatics pipeline for base-calling, contig assembly, quality trimming, immunogenetic annotation with Cloanalyst (https://www.bu.edu/computationalimmunology/research/software/), VDJ sequence quality filtering, functionality assessment, and isotyping as described^22^.

For generation of monoclonal antibodies bearing BCR from antigen-reactive B cells of interest, commercially-obtained plasmids (GeneScript, Piscataway, NJ) with antibody heavy and light chain genes were used to transfect suspension Expi 293i cells using ExpiFectamine 293 transfection reagents (Life Technologies, Gibco; Cat#A14524) as described^8,21,23^. Purified recombinant mAbs were dialyzed against PBS, analyzed, and stored at −4°C^8,23,24^. All recombinant mAbs were expressed from plasmids encoding a human IgG constant region, and were QC’ed in Western Blot for appropriate heavy and light chain protein expression and/or via size exclusion chromatography (SEC) for a single peak indicative of stable antibody expression.

### Single cell immune profiling assay (10X Genomics)

We performed single cell immune profiling assay (10X Genomics) to generate paired variable heavy and light chain genes as well as transcriptome sequences from single CH505TF gp120-reactive B cells that were sorted via flow cytometry. Total PBMCs were stained using antibody cocktails to facilitate flow sorting of B cells as described^21,23^. The antibody cocktails also included CH505TF gp120 conjugated to AF647 and VB515 fluorophores to facilitate the detection of antigen-reactive B cells as previously described^8^.

Bulk sorted antigen-reactive B cells were subjected to single cell immune profiling assays for BCR and transcriptome sequencing and analysis as described^21,22^. Briefly, single cell VDJ library construction was performed using 10X Chromium Controller using the Chromium Next GEM Single Cell 5’ and BCR amplification kits, and sequenced via Illumina sequencing platforms. BCR analysis was performed using the Cell Ranger single cell gene expression software provided by 10X Genomics as described^21^. For analysis of transcriptome sequences, the cell ranger software suite was used to generate sequencing fastq files and to perform sample de-multiplexing, barcode processing, reference alignment and single-cell 3’ gene counting as described^22^. All participants studied received five vaccinations except participant 115-42 who received vaccines 1-3 and was subsequently lost to follow-up. We studied BCR repertoire analysis post final vaccination in all the vaccinees; post 5^th^ or post 3^rd^ (for participant 115-42 only).

### Negative stain electron microscopy (NSEM)

MAb was diluted to 100 µg/ml with 5% Gly-HBS buffer containing 8 mM glutaraldehyde. After 5 minute incubation, glutaraldehyde was quenched by adding sufficient 1 M Tris stock, pH 7.4, to give 80 mM final Tris concentration and incubated for 5 minutes. Quenched sample was applied to a glow-discharged carbon-coated EM grid for 10-12 seconds, then blotted, and stained with 2 g/dL uranyl formate for 1 minute, blotted and air-dried. Grids were examined on a Philips EM420 electron microscope operating at 120 kV and nominal magnification of 49,000x, and 104 images were collected on a 76 Mpix CCD camera at 2.4 Å/pixel. Images were analyzed by 2D class averages and 3D reconstructions calculated using standard protocols with Relion 3.0^25^.

### Initial Processing of Single-Cell RNA-seq Data

#### Raw Data Processing

Raw single-cell RNA sequencing data were processed using Cell Ranger (v3.1.0, 10x Genomics)^26^. For each sample, FASTQ files were generated with the cellranger mkfastq command and vdj pipeline was used for quantifying cell count with GRCh38 reference (refdata-cellranger-vdj-GRCh38-alts-ensembl-3.1.0).

#### Data Import and Seurat Object Creation

Count matrices were imported into R (v4.0.2) using Seurat (v3.1.0)^27^. Each dataset was converted into a Seurat object with the following parameters: a minimum of 5 cells per gene (min.cells = 5) and at least 200 detected genes per cell (min.features = 200). The percentage of mitochondrial transcripts per cell was computed using the PercentageFeatureSet() function in Seurat.

#### Quality Control and Filtering

Cells were filtered on a per-sample basis to exclude low-quality cells and potential doublets. Filtering thresholds were defined using the number of detected genes (nFeature_RNA), UMI counts (nCount_RNA), and percentage of mitochondrial reads (percent.mt) as listed for each vaccine trial participant (vaccine (V) and placebo (P) recipients) studied: <u>V1</u>: nGene < 4000, nUMI < 15,000, percent.mt < 20%; <u>V2</u>: nGene < 6000, nUMI < 40,000, percent.mt < 20%; <u>V3</u>: nGene < 4000, nUMI < 20,000, percent.mt < 20%; <u>V4</u>: nGene < 4000, nUMI < 20,000, percent.mt < 20%; <u>V5</u>: nGene < 6000, nUMI < 40,000, percent.mt < 20%; <u>V6</u>: nGene < 4000, nUMI < 20,000, percent.mt < 30%; <u>V7</u>: nGene < 4000, nUMI < 20,000, percent.mt < 20%; <u>V8</u>: nGene < 6000, nUMI < 50,000, percent.mt < 30%; <u>V9</u>: nGene < 6000, nUMI < 40,000, percent.mt < 30%; <u>V10</u>: nGene < 2000, nUMI < 8000, percent.mt < 40%; <u>V11</u>: nGene < 3500, nUMI < 15,000, percent.mt < 30%; <u>V12</u>: nGene < 6000, nUMI < 50,000, percent.mt < 30%; <u>P1</u>: nGene < 4000, nUMI < 20,000, percent.mt < 20%; <u>P2</u>: nGene < 3500, nUMI < 20,000, percent.mt < 30%; and <u>P3</u>: nGene < 4000, nUMI < 20,000, percent.mt < 20%. Cells outside these thresholds were excluded from downstream analyses.

#### Normalization and Feature Selection

Filtered data were normalized using log normalization (NormalizeData with scale factor = 10,000). Highly variable features were identified using the vst method (FindVariableFeatures, n = 2000). These processed Seurat objects were subsequently used for downstream integration and clustering analyses.

#### Data Integration

After quality control and normalization, individual Seurat objects for each sample (V1– V12, P1–P3) were compiled into a list and merged. Datasets were integrated using the Seurat v3 workflow. Briefly, integration anchors were identified with FindIntegrationAnchors (dims = 1:30), followed by dataset integration with IntegrateData across the first 30 dimensions. The integrated object was scaled with ScaleData, and principal component analysis (PCA) was performed on 100 components using the top 2000 variable features.

#### Dimensionality Reduction and Clustering

The number of informative PCs was assessed by inspection of PC heatmaps, JackStraw resampling, and elbow plots. Based on these diagnostics, the first 21 PCs were selected for downstream clustering and visualization. Graph-based clustering was performed using FindNeighbors and FindClusters across multiple resolutions (0.4–1.2), with 0.8 chosen as the final resolution based on clustree analysis of cluster stability. Nonlinear dimensionality reduction was performed using UMAP (RunUMAP) on the first 21 PCs.

#### Visualization

Cells were visualized using Seurat’s DimPlot function, with clusters labeled on UMAP embeddings. Additional visualizations included sample-wise and condition-wise splits (group.by = “library” and split.by = “orig.ident”), enabling assessment of batch effects and sample-specific distributions.

#### Cell Type Annotation

Automated cell type annotation was performed using SingleR (v1.4.1)^28^ with the Monaco Immune reference dataset. Both *main* (broad) and *fine* (subtype) immune labels were assigned to each cluster, and visualized on UMAP projections. Annotation results were stored in the Seurat object metadata for downstream interpretation.

#### Cluster Cell Frequencies

Cluster composition across libraries (samples/conditions) was quantified by tabulating cell counts per cluster per library. Counts and percentages were exported as CSV files for statistical analysis and visualization.

#### Clonotype-Specific Subsetting

To investigate the distribution of immunoglobulin heavy chain variable (IGHV) gene clonotypes, cells expressing IGHV1-2, IGHV1-46, and IGHV4-59 were identified from V(D)J assignments and extracted as subsets. Seurat objects were created for each clonotype, and merged with the remaining (“unselected”) cells to compare clonotype-specific versus background B cell populations.

## Statistics

### BAMA

Several criteria were used to determine if data from an assay were acceptable and could be statistically analyzed. First, the blood draw date must have been within the allowable visit window as determined by the protocol. Second, if the blank bead negative control exceeded 5,000 MFI, the sample was repeated. If the repeat value exceeded 5,000 MFI, the sample was excluded from analysis due to high background.

The preset assay criteria for sample reporting were coefficient of variation (CV) per duplicate values for each sample is <15% and >100 beads counted per sample. To control for protein performance, the positive control titer included on each assay must be within +/-3 standard deviations of the mean for each antigen (tracked with a Levey-Jennings plot with preset acceptance of titer and calculated with a four-parameter logistic equation).

### ICS

To assess positivity for a peptide pool within a T-cell subset, a two-by-two contingency table was constructed comparing the HIV-1 peptide stimulated and negative control data. The four entries in each table were the number of cells positive for IL-2 and/or IFN-γ (IFN-γ or IL-2 or CD40L) and the number of cells negative for IL-2 and/or IFN-γ (IFN-γ or IL-2 or CD40L), for both the stimulated and the negative control data. If both negative control replicates were included, then the average number of total cells and the average number of positive cells were used. A one-sided Fisher’s exact test was applied to the table, testing whether the number of cytokine-producing cells for the stimulated data was equal to that for the negative control data. Since multiple individual tests (for each peptide pool) were conducted simultaneously, a multiplicity adjustment was made to the individual peptide pool p-values using the Bonferroni-Holm adjustment method. If the adjusted p-value for a peptide pool was ≤0.00001, the response to the peptide pool for the T-cell subset was considered positive. Because the sample sizes (i.e., total cell counts for the T-cell subset) were large, e.g., as high as 100,000 cells, the Fisher’s exact test had high power to reject the null hypothesis for very small differences. Therefore, the adjusted p-value significance threshold was chosen stringently (≤0.00001). If at least one peptide pool for a specific HIV-1 protein was positive, then the overall response to the protein was considered positive. If any peptide pool was positive for a T-cell subset, then the overall response for that T-cell subset was considered positive. Pooling of the Env peptides was included: any Env PTEg: sum of magnitude of response to Env-1-PTEg-SEQ, Env-2-PTEg-SEQ.

### nAb responses

Response to a virus/isolate in the TZM-bl assay was considered positive if the neutralization titer was above a pre-specified cutoff (one-half the lowest dilution tested). A titer was defined as the serum dilution that reduces RLUs by 50%, relative to the RLUs in virus control wells (cells + virus only), after subtraction of background RLU (cells only). The pre-specified cutoff was 10 for TZM-bl cells. Response rates and corresponding 95% confidence intervals were calculated by the Wilson score method^29^. Descriptive statistics were summarized by treatment arm for response magnitudes among all participants and among positive responders only. The Kruskal-Wallis rank test was performed to compare the magnitudes among all the vaccine groups. The Wilcoxon rank sum test was used to compare magnitudes between pairs of adjacent vaccine arms (T1 vs T2 and T2 vs T3). Hodges-Lehmann point estimates and 2-sided (1-0.05/k) × 100% confidence intervals (CIs) were constructed for the differences in location centers of the k = 2 pairwise comparisons of vaccine arms. Pooled response rates and response magnitudes were compared across isolates (CH0505s.G458Y.N279K.2/GnT1-vs. CH0505s.G458Y.N279K.N280D.6/GnT1-) using McNemar’s test and a Wilcoxon signed-rank test, respectively.

## RESULTS

### Study design and participant demographics

This multi-site randomized placebo-controlled trial was conducted in two parts (**Figure 1**), with Part A assessing the safety and immunogenicity of increasing doses (20, 100, 400 mcg) of the recombinant CH505 transmitted founder gp120 protein (CH505wTF) subunit vaccine in combination with the GLA-SE adjuvant (10 mcg). Informed by Part A, the dose of gp120 proteins (CH505wTF, CH505w53, and CH505w78) (400 mcg) was selected for Part B. The objectives of Part B were to assess the safety and immunogenicity of sequential or additive vaccination with CH103 lineage optimized gp120 proteins, and to evaluate repeated vaccination with the CH505 M5 gp120 protein, intended to elicit bnAb precursors to another lineage, CH235.

Forty-two participants were enrolled in Part A between October 2017 and May 2018 (**Figure 1 and Supplemental Figure 1)**. Part B enrolled sixty-five participants from December 2020 to October 2022. In total, 59.5% were assigned male sex at birth, 2.4% identified as Hispanic or Latino, and 65.7% were White (**Table 1**). The median age of participants was 29 years (range 19-49).

**Table 1.** HVTN 115 participant demographics.

| Characteristic | T1 (N = 12)<sup>1</sup> | T2 (N = 12)<sup>1</sup> | T3 (N = 12)<sup>1</sup> | C4 (N = 6)<sup>1</sup> | T5 (N = 20)<sup>1</sup> | T6 (N = 20)<sup>1</sup> | T7 (N = 20)<sup>1</sup> |
| --- | --- | --- | --- | --- | --- | --- | --- |
| **Sex** |  |  |  |  |  |  |  |
| Male | 7 (58%) | 7 (58%) | 8 (67%) | 3 (50%) | 13 (65%) | 11 (55%) | 12 (60%) |
| Female | 5 (42%) | 5 (42%) | 4 (33%) | 3 (50%) | 7 (35%) | 9 (45%) | 8 (40%) |
| **Ethnicity** |  |  |  |  |  |  |  |
| Hispanic or Latino | 0 (0%) | 1 (8.3%) | 0 (0%) | 0 (0%) | 3 (15%) | 3 (15%) | 5 (25%) |
| Not Hispanic or Latino | 12 (100%) | 11 (92%) | 12 (100%) | 6 (100%) | 17 (85%) | 17 (85%) | 15 (75%) |
| **Race** |  |  |  |  |  |  |  |
| Native Hawaiian or other Pacific Islander | 0 (0%) | 0 (0%) | 0 (0%) | 0 (0%) | 0 (0%) | 0 (0%) | 1 (5.0%) |
| White | 8 (67%) | 7 (58%) | 6 (50%) | 4 (67%) | 15 (75%) | 15 (75%) | 12 (60%) |
| Black or African American | 3 (25%) | 1 (8.3%) | 3 (25%) | 1 (17%) | 1 (5.0%) | 1 (5.0%) | 3 (15%) |
| Asian | 0 (0%) | 1 (8.3%) | 3 (25%) | 0 (0%) | 2 (10%) | 0 (0%) | 2 (10%) |
| American Indian or Alaskan Native | 0 (0%) | 0 (0%) | 0 (0%) | 0 (0%) | 1 (5.0%) | 0 (0%) | 0 (0%) |
| Multiracial | 1 (8.3%) | 2 (17%) | 0 (0%) | 1 (17%) | 0 (0%) | 3 (15%) | 1 (5.0%) |
| Other | 0 (0%) | 1 (8.3%) | 0 (0%) | 0 (0%) | 1 (5.0%) | 1 (5.0%) | 1 (5.0%) |
| **Age (Years)** |  |  |  |  |  |  |  |
| Less than 18 | 0 (0%) | 0 (0%) | 0 (0%) | 0 (0%) | 0 (0%) | 0 (0%) | 0 (0%) |
| 18 - 20 | 1 (8.3%) | 1 (8.3%) | 1 (8.3%) | 1 (17%) | 0 (0%) | 2 (10%) | 0 (0%) |
| 21 - 30 | 4 (33%) | 5 (42%) | 7 (58%) | 4 (67%) | 12 (60%) | 11 (55%) | 11 (55%) |
| 31 - 40 | 5 (42%) | 4 (33%) | 2 (17%) | 0 (0%) | 6 (30%) | 5 (25%) | 7 (35%) |
| 41 - 50 | 2 (17%) | 2 (17%) | 2 (17%) | 1 (17%) | 2 (10%) | 2 (10%) | 2 (10%) |
| Over 50 | 0 (0%) | 0 (0%) | 0 (0%) | 0 (0%) | 0 (0%) | 0 (0%) | 0 (0%) |
| Median | 32 | 31 | 27 | 28 | 29 | 28 | 28 |
| Range | 20 - 48 | 19 - 49 | 20 - 48 | 19 - 45 | 21 - 43 | 18 - 42 | 22 - 48 |
| Day 0/Enrollment | 12 (100%) | 12 (100%) | 12 (100%) | 6 (100%) | 20 (100%) | 20 (100%) | 20 (100%) |
| Day 56 | 10 (83%) | 11 (92%) | 12 (100%) | 5 (83%) | 20 (100%) | 19 (95%) | 18 (90%) |
| Day 112 | 10 (83%) | 10 (83%) | 12 (100%) | 4 (67%) | 18 (90%) | 20 (100%) | 19 (95%) |
| Day 224 | 9 (75%) | 10 (83%) | 11 (92%) | 4 (67%) | 17 (85%) | 19 (95%) | 19 (95%) |
| Day 364 | 9 (75%) | 9 (75%) | 11 (92%) | 4 (67%) | - | - | - |

<sup>1</sup>n (%); Median; Min - Max

### Safety and Tolerability

Overall, the vaccines were found to be safe and well-tolerated by participants. Most solicited adverse events (“reactogenicity”) were mild to moderate, and all resolved within the seven-day observation period, with most resolving within 72 hours. No significant difference in the rate of solicited adverse events was observed when comparing the first administration to other vaccinations.

There were two Part A participants (one in T2 and one in T3) and six Part B participants with grade 3 erythema/redness and induration/swelling (**Figure 2A, C)**. Of note, the Grade 3 erythema did not interfere with daily activities, and the rate of erythema was not obviously different by dose or product. For systemic solicited adverse events, there were two participants in Part A with grade 3 headache and four participants in Part B with transient grade 3 malaise/fatigue (**Figure 2B, D**). Among the unsolicited adverse events deemed related to study product of grade 2 or greater, there was one grade 2 injection site cellulitis in a T2 participant in part A, and one participant in Part B with two days transient knee joint pain shortly after the second vaccination. Transient grade 1 adverse events deemed related to study product included injection site pruritis (7 participants), lymphadenopathy (one participant) elevated AST (1 participants). There were no SAEs deemed related to study product during the trial. Two participants in Part A discontinued vaccination because they refused further vaccinations (one in the 100 mcg and one in the 400 mcg dose group), and one participant in Part B discontinued further vaccinations due to solicited adverse events.

**Figure 2.**
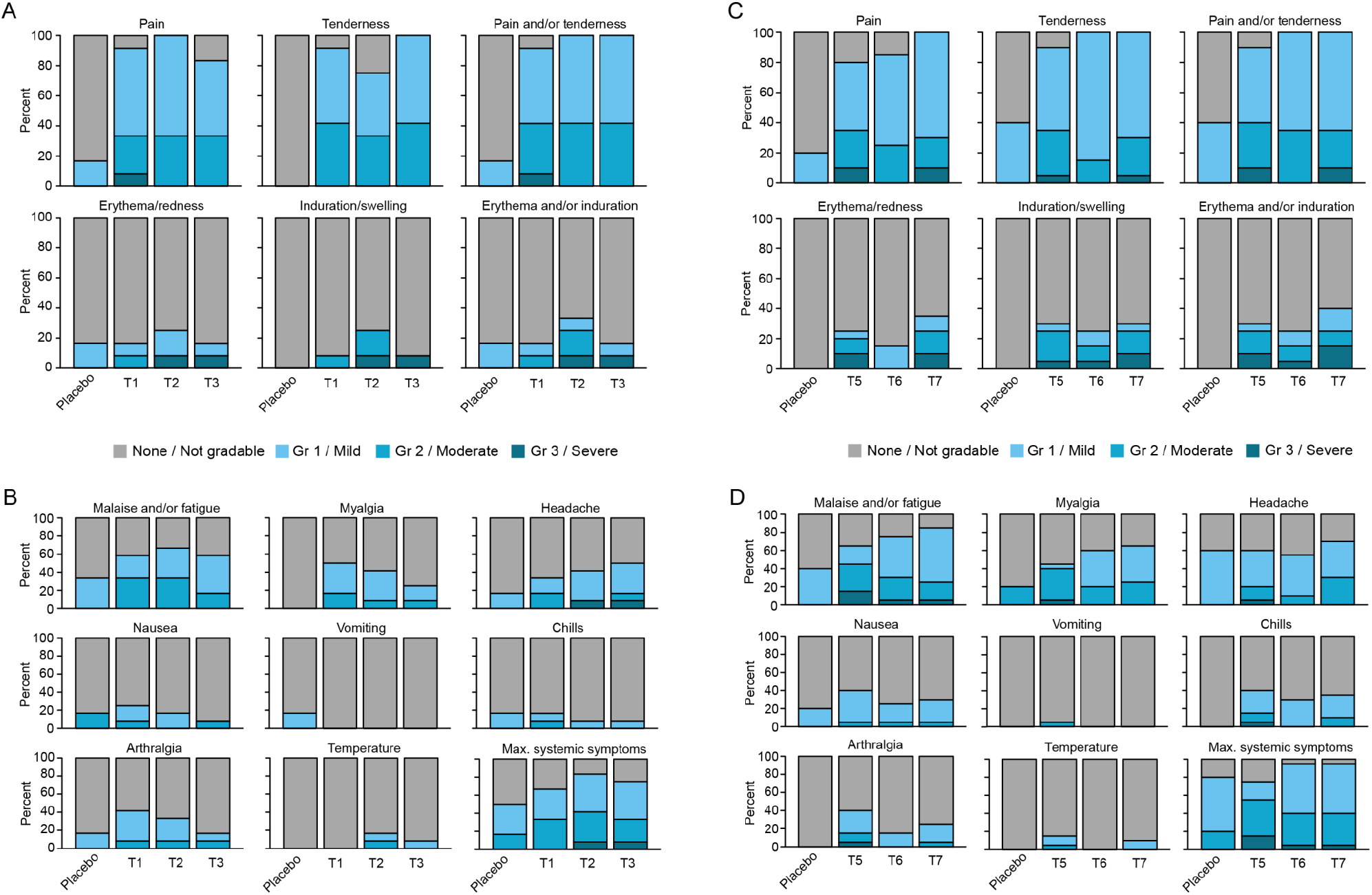
Safety and reactogenicity. **A)** Local reactogenicity for Part A. **B)** Systemic reactogenicity for Part A. **C)** Local reactogenicity for Part B. **D)** Systemic reactogenicity for Part B.

### CH505TF immunogenicity (Part A)

At two weeks after the third vaccination (M4.5), response rates of Env-specific CD4+ T cells, as defined by expression of IFNγ, IL2, or CD40L, were 67-90%, with no significant differences detected among the vaccine arms in response magnitudes or response rates (**Figure 3A**). At two weeks after the final vaccination (M12.5), the Env-specific CD4+ T cell positive response rate were 36-56%, with magnitudes significantly decreased in all vaccine arms compared to M4.5 (T1: 0.146 vs 0.121, p=0.0078; T2: 0.231 vs 0.071, p=0.0156; T3: 0.237 vs 0.085, p=0.0137), with no significant differences detected among the vaccine arms in response magnitudes or response rates at M12.5. The Env-specific CD4+ T cell response was largely confined to cells expressing CD40L, but not IFNγ or IL2 (not shown). For CD8+ T cells, no positive responses were observed (not shown).

**Figure 3.**
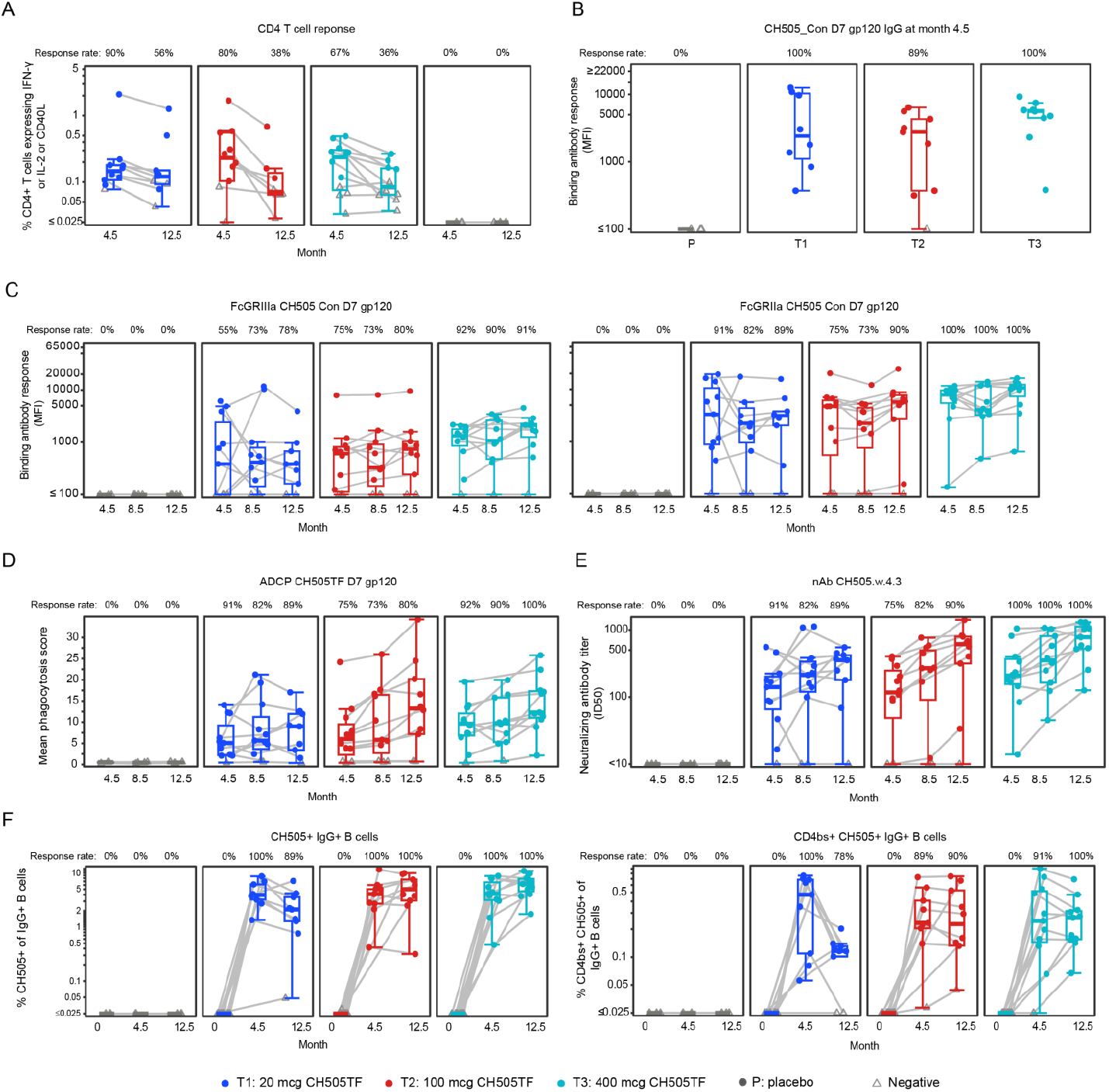
Part A immunogenicity. **A)** Peripheral blood CD4+ response to any Env peptide pool. **B)** Serum binding IgG response against CH505 Con_D7 gp120. **C)** Serum FcγRIIIa F158 and FcγRIIa H131 binding antibody response to CH505 Con_D7 gp120. **D)** Serum ADCP of CH505TF D7 gp120 coated beads. **E)** Serum neutralizing antibody titers against CH505.w.4.3. **F)** Peripheral blood CH505 positive and CH505 CH505 positive CD4bs positive IgG+ B cells. Each point represents the magnitude of response for an individual vaccine recipient at each time point. Boxplots summarize the distribution of responses. Gray lines connect observations from the same study participant. Filled circles indicate positive responses, gray open triangles indicate nonresponders. Response rates for individual groups are noted above each boxplot.

At two weeks after the third vaccination (M4.5), there were response rates of serum IgG binding antibody to the autologous monomeric CH505TF gp120 of 89% (T2) - 100% (T1+T3) (**Figure 3B**), with 100% response rate achieved in all vaccine arms by two weeks after the final vaccination (M12.5) (not shown). Only one participant, in T3 developed low IgG binding antibody titers to the SOSIP trimeric CH505TF (not shown). At two weeks after the third vaccination, the response rate of FcGRIIIa binding IgG to CH505TF gp120 was highest in T3 (92%), with the response rates increasing further in T1 and T2 with subsequent vaccinations, reaching 78% and 80% respectively two weeks after the final vaccination (**Figure 3C**). The response rates of FcGRIIa binding IgG to CH505TF gp120 similarly were highest in T3 (92-100%) at all timepoints. Two weeks after the third vaccination (M4.5), response rates of antibody-dependent cellular phagocytosis (ADCP) of CH505TF gp120 coated beads were high in all vaccine arms (75-92%), with 100% response rate in T3 two weeks after the final vaccination (M12.5) (**Figure 3D**). At two weeks after the third vaccination, the response rates of autologous neutralizing antibody to Tier 1A virus (CH505.w4.3) were 91%, 75%, and 100% in T1, T2, and T3 respectively (**Figure 3E**), and at two weeks after the final vaccination the response rates were 89%, 90%, and 100% in T1, T2, and T3 respectively. In general, the titers of neutralizing antibody increased with subsequent vaccinations. No autologous Tier 2 virus neutralization was detected (not shown).

The induction of CH505TF gp120 and CD4 binding site (CD4bs)-specific B cells was measured by flow cytometry. At baseline (M0), the frequencies of the total (not shown) and IgG+ CH505TF gp120 and CD4bs-specific B cells (**Figure 3F**) were close to zero among all treatment arms. At two weeks after the third immunization (M4.5), all participants in vaccine arms had detectable CH505TF gp120 IgG+ B cell responses, with 3.90%, 4.07%, and 4.14% group median CH505TF gp120 IgG+ B cells in T1, T2 and T3 respectively, with no significant differences detected between vaccine arms. At this time, response rates were 100%, 89%, and 91%, with 0.47%, 0.23%, and 0.24% group median CH505TF CD4bs IgG+ B cells in T1, T2, and T3 respectively, with no significant differences detected between arms. At two weeks after the final immunization (M12.5), response rates were 89%, 100%, and 100%, with 2.12%, 5.01%, and 6.27% CH505TF gp120 IgG+ B cells in T1, T2 and T3 respectively, with T1 being significantly lower than T2 (p=0.0350) and T3 (p=0.0042). At this time, response rates were 78%, 90%, and 100%, with 0.11%, 0.23%, and 0.27% CH505TF CD4bs IgG+ B cells in T1, T2, and T3 respectively, with T1 being significantly lower than T2 (p=0.0076) and T3 (p=0.0023). The frequency of CH505TF gp120 IgG+ and CH505TF CD4bs IgG+ B cells did not significantly change in T2 and T3 between the third and final immunization, however, their frequencies declined for T1 with the decrease in CH505TF gp120 CD4bs IgG+ B cells reaching significance (p=0.0391). These results indicate that CH505TF gp120 induced CD4bs-specific IgG+ B cells, with highest response rate observed with the 400 mcg dose.

### Sequential CH505 gp120 immunogenicity (Part B)

Based on the immunogenicity results of CH505TF gp120 alone (Part A), particularly the 100% response rate of autologous neutralizing antibody after 3 vaccinations, the 400 mcg dose was chosen for evaluation of vaccination with sequential (T5, “seq”) or additive (T6, “add”) vaccinations with CH505 gp120 proteins or repeated vaccination with CH505 M5 gp120 protein (T7) (**Figure 1**).

Two weeks following the second vaccination (M2.5), nearly all participants (95-100% response rates) developed IgG binding antibody to CH505TF, CH505 M5, CH505.w53, and CH505.w78 gp120 (**Figure 4A**). To assess potential CD4bs–targeting IgG responses, IgG binding antibody to CD4bs mutated CH505 gp120 proteins were also evaluated. CD4bs specific antibodies identified by differential binding profiles were elicited in all immunized participants following the second vaccination with response rates and magnitudes comparable between T5 (seq) and T6 (add). A trend toward lower IgG titers to CD4bs mutants in T5 (seq) was observed. However, a significant reduction in the number of CD4bs positive responders following the fourth vaccination (M8.5) were observed across all immunized groups (**Figure 4B,C**), suggesting that repetitive immunization with the CH505 gp120 immunogens may promote the development of antibodies targeting epitopes outside the CD4bs.

**Figure 4.**
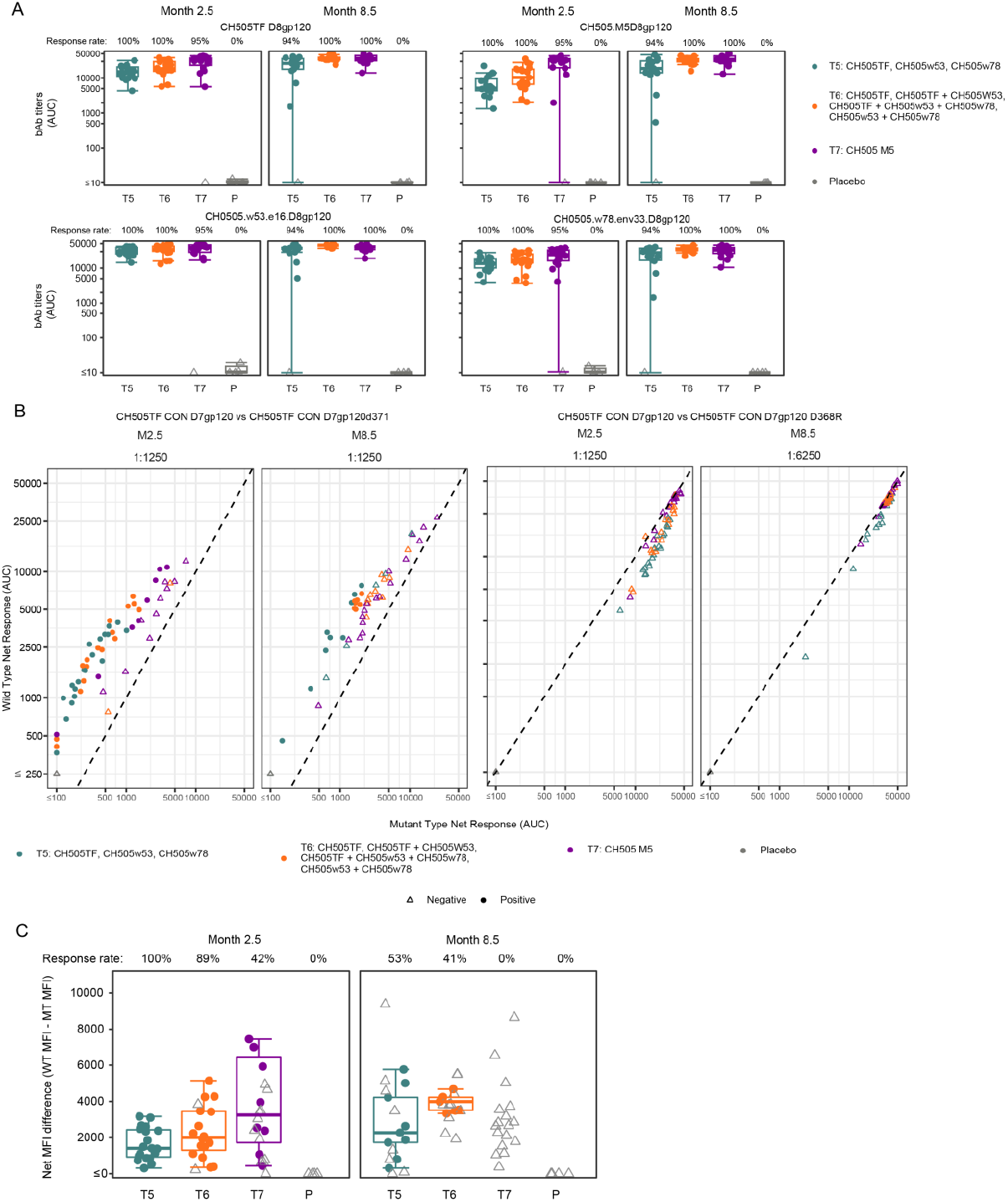
Part B HIV Env CD4 binding site serum IgG response. Serum binding IgG response against CH505TF D7 gp120 (WT) and CD4bs d371 and D368R gp120 mutants. **A)** Response to individual gp120s. Each point represents the magnitude of response (AUC) for an individual vaccine recipient at each time point. Boxplots summarize the distribution of responses. Gray lines connect observations from the same study participant. Filled circles indicate positive responses, gray open triangles indicate nonresponders. Response rates for individual groups are noted above each boxplot. **B)** Differential binding profiles of WT vs. CD4bs mutant proteins. **C)** Derived CD4bs differential binding response magnitude and response rate WT vs CD4bs d371 mutant for M2.5 and M8.5.

Vaccination with the CH505 M5 (T7) resulted in the greatest V1V2 IgG response rates and titers after the second and final vaccinations (**Figure 5A**), with T6 (add) having significantly higher response rates and magnitude in V1V2 IgG compared to T5 (seq). These results suggest the sequential immunization strategy may slightly favor CD4bs antibody development, and the additive immunization strategy favor V1V2 antibody development.

**Figure 5.**
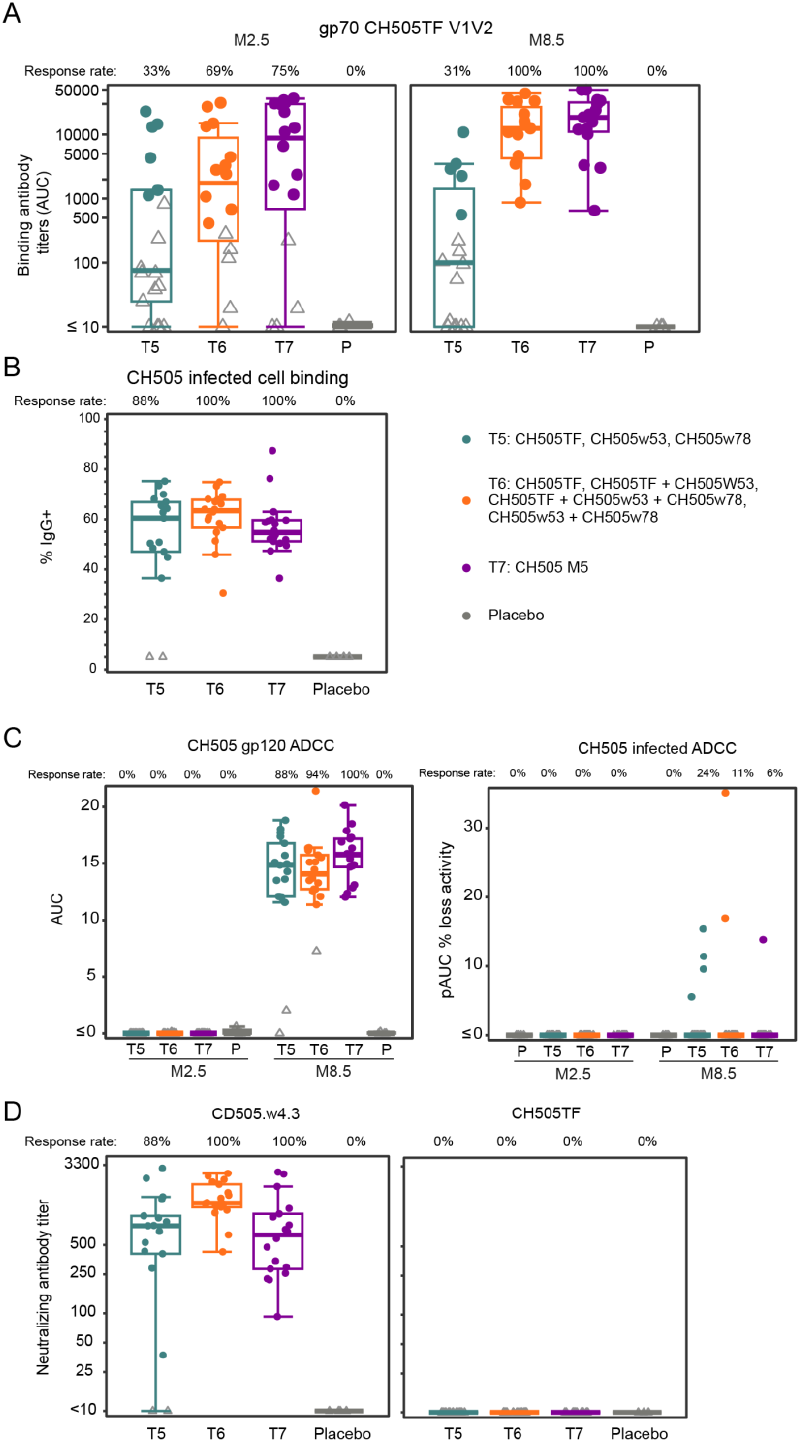
Part B immunogenicity. **A)** Serum binding IgG response against CH505 V1V2. **B)** Serum IgG binding response against CEM.CCR5.NKR cells infected with CH505. **C)** Serum ADCC response against CH505TF D7gp120 coated target cells (GranToxiLux) or CH0505s.LucR.T2A.ecto/293T/17 infected target cells. **D)** Serum neutralizing antibody titers against CH505.w.4.3, CH505TF.G458Y.N279K.2, and CH505TF.gly4. Each point represents the magnitude of response for an individual vaccine recipient at each time point. Boxplots summarize the distribution of responses. Filled symbols indicate positive responses, gray open symbols indicate nonresponders. Response rates for individual groups are noted above each boxplot.

At two weeks after the final vaccination (M8.5), all vaccine arms had similar high response rates (88-100%) of IgG serum antibodies binding to cells infected with CH505, with 52%, 61%, and 57% IgG+ bound cells in T5, T6, and T7 respectively (**Figure 5B**). All vaccine arms had similar antibody-dependent cellular cytotoxicity (ADCC) response rates to target cells coated with CH505TF gp120 (88%-100%), however lower response rates to CH505 infected target cells (6%-24%) (**Figure 5C**). At two weeks after the final vaccination, the response rates of autologous neutralizing antibody to Tier 1A virus (CH505.w4.3) were 88%, 100%, and 100% in T5, T6, and T7 respectively, with the highest titer observed in T6 (**Figure 5D**). No neutralization was detected against the autologous tier 2 or difficult-to-neutralize vaccine strain viruses in any group (not shown), in contrast with high neutralization titers against tier 1 or easy-to-neutralize vaccine strain CH505.w4.3 (**Figure 5D**). Overall, these results indicate that, while immunogenic, the sequential CH505 gp120 proteins (T5+T6) and the CH505 M5 gp120 protein induced neutralizing antibodies with limited breadth.

### Vaccine-induced B cell responses

Observing the limited induction of neutralizing antibody breadth in Part B, we sought to further define the features of the B cells induced by CH505TF gp120 vaccination, to potentially discern the basis of this outcome. CH505TF gp120 antigen-reactive B cells were sorted by flow cytometry from all twelve vaccinees post-fifth or final immunization from Part A in T3 (**Figure 6A**). For comparison, we sorted the antigen-negative B cells from three placebo recipients in T4 as no antigen-reactive B cells were identified in placebo recipients (**Suppl Figure 3**). Flow sorted B cell populations were subjected to 10X genomics single cell immune profiling assay that generated paired heavy and light chain genes and transcriptome sequences from single antigen-sorted B cells (**Supp Table 4 and Suppl Figure 4)**. Single cell transcriptional analysis of sorted cell populations revealed 13 transcriptionally distinct clusters (**Figure 6B**). Five unique gene expression clusters (0, 2, 8, 10, 11) were observed in the vaccine recipients (**Figure 6C; Suppl Figure 4A**). Using singleR annotation with the Monaco Immune reference dataset (see Methods), these unique clusters in the vaccine recipients were transcriptionally defined as switched memory B cell populations (**Suppl Table 4**); a hallmark phenotype of antigen-specific vaccine-induced B cells

**Figure 6:**
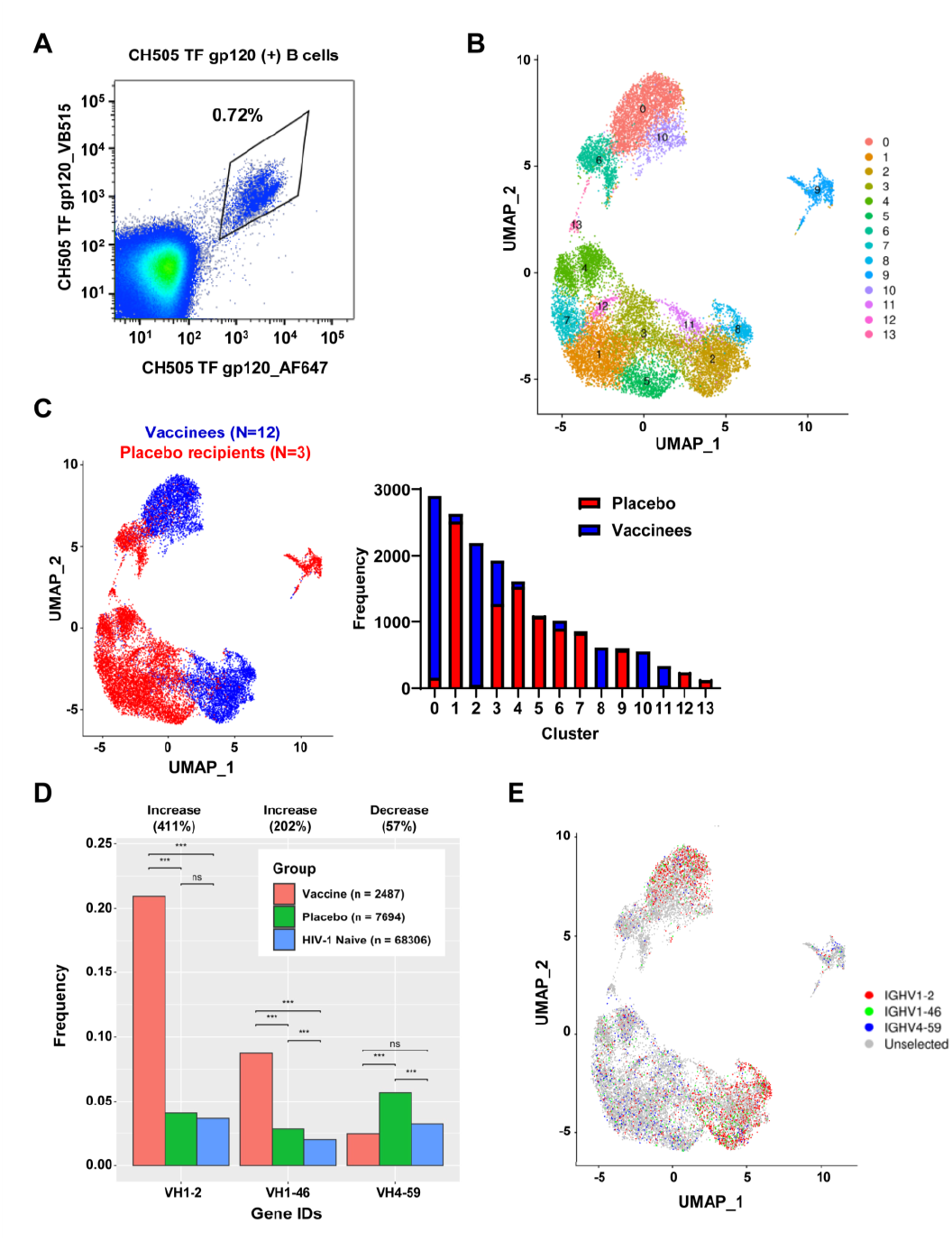
CH505 TF gp120 induced B cell response. **A)** Representative flow cytometry gating for sorting of CH505 TF gp120+ B cells post-fifth immunization. Analysis of CH505 TF gp120+ B cells from vaccinees post-fifth immunization (N=12) and antigen-negative B cells from placebo recipients (N=3) subject to 10X genomics single cell immune profiling assay. **B)** UMAP defining 13 transcriptionally unique antigen-reactive B cell clusters. **C)** UMAP and bar graph comparing the frequency of antigen-reactive B cells from vaccinees and placebo recipients within the 13 unique clusters. **D)** Frequency of VH gene usage in antigen-reactive B cells in vaccinees (N=12), placebo (N=3), and HIV-1 naïve (N=3) individuals. **E)** UMAP of antigen-reactive VH gene usage. P-value = *>0.05, **>0.01, ***>0.001, ****>0.0001.

Additionally, we found that antigen-reactive B cells used a wide range of variable heavy chain (VH) genes and had unique immunogenetics compared to non-antigen-reactive B cells. In particular, antigen-reactive B cells had an increase in length (amino acids) of the VH third complementarity determining region (CDR3H), increased VH somatic hypermutation (SHM) frequency, and antibody isotype switching to IGHG1, relative to antigen-negative B cells in placebo participants (**Suppl Figure 5**).

We focused our analysis on the canonical CD4-binding site (BS) antibody heavy chain genes used by known bnAbs VRC01 (VH1-2), CH235 (VH1-46), and CH103 (VH4-59). Frequencies of antigen-reactive B cells in vaccine recipients that used VH1-2 and VH1-46 genes were increased 411% and 202%, respectively, compared to antigen-negative B cells from the placebo participants (**Figure 6D, Suppl Figure 5A**). Moreover, the VH1-2 and VH1-46 genes were predominantly located in the unique vaccinee gene expression clusters defined as switched memory B cells, indicating a vaccine-induced expansion of these cell subsets (**Figure 6E**). Dominant expression of VH1-2 prompted us to investigate if there was a vaccine-driven expansion of VRC01-like B cells. We observed no VH1-2 clones paired with the VRC01-signature short CDR3 light chain of 5 amino acids (data not shown). Taken together, these results highlight the induction of transcriptionally distinct isotype switch memory B cells displaying a B cell repertoire post vaccination with similar immunogenetics used by canonical HIV-1 Env-reactive CD4BS antibodies.

Next, we investigated whether the CH505TF gp120 elicited CD4BS bnAb precursors in the twelve vaccine recipients. We used a three-color antigen-specific flow cytometry sorting strategy designed to isolate vaccine-induced CH103-like CD4BS bnAb precursor B cells in twelve vaccinees, post third immunization (**Figure 7A, Suppl Figure 6**). Here, we sorted CD20+ B cells that reacted to wild-type CH505TF SOSIP trimer conjugated to two different fluorophores (AF647 and VB515), but did not bind STG (S365K, T445E, G459E)-mutant^30^ of CH505TF SOSIP with a disruption of the CD4BS that was conjugated to BV421 fluorophore. We sorted a total of 337 B cells, from which we recovered heavy and light chain genes from 172 B cells. The paired heavy and light chain genes recovered from each B cell were used to generate recombinant monoclonal antibodies (mAbs) to test for binding specificities and function.

**Figure 7:**
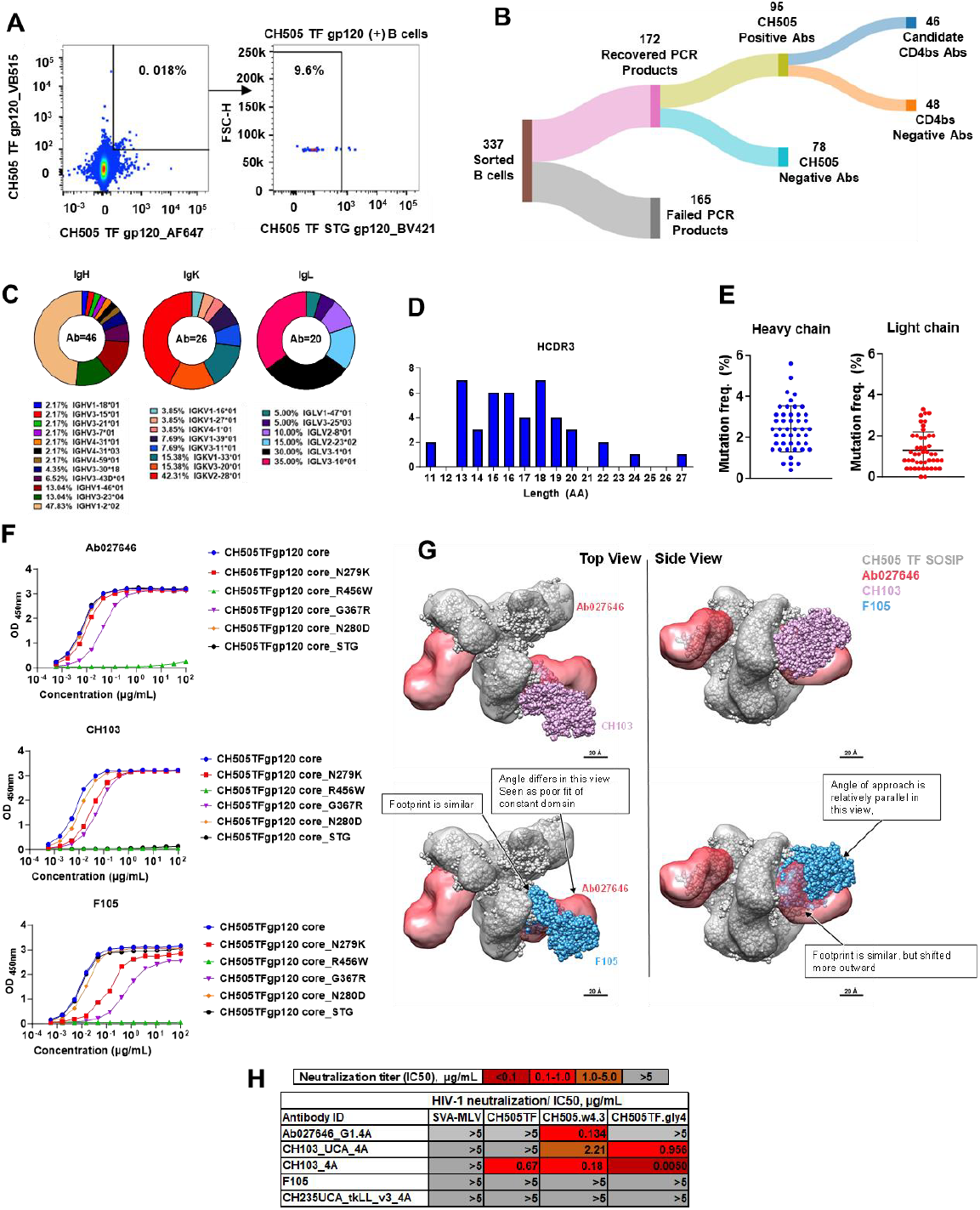
CH505 TF gp120 induced mAb profiling. **A)** Representative flow cytometry gating for sorting of antigen-reactive CH505 TF gp120+ CH505 TF STG-B cells for single cell PCR from vaccinees post-third immunization (N=12). **B)** Isolation and testing of antigen-reactive CH505 TF gp120+ CH505 TF STG-B cell VDJ sequences to determine candidate CD4bs Abs. **C)** IgH, IgK, and IgL immunogenetics of candidate CD4bs Abs (N=46). **D)** HCDR3 length of candidate CD4bs Abs (N=46). **E)** Heavy and light chain mutation frequency of candidate CD4bs Abs (N=46). **F)** ELISA of Ab027646, CH103, and F105 for reactivity to the core epitope of gp120 and various mutant peptides. Reflective of multiple experiments. **G)** NSEM of CH103 (pink) or F105 (blue) binding footprint compared to Ab027646 (red) in complex with CH505 TF SOSIP (grey). **H**) TZM-bl neutralization assay of Tier 1 and Tier 2 HIV-1 viruses or control SVA-MLV.

Of the 172 mAbs, 94 proved to be CH505 Env-reactive, with 46 of these antibodies from six vaccinees [115-04 (n=2), 115-17 (n=5), 115-27 (n=3), 115-30 (n=1), 115-41 (n=32) and 115-60 (n=3)] determined to be candidate CD4BS antibodies based on antigen binding profiles (**Figure 7B; Suppl Table 5**). The heavy and light chain genes from the candidate CD4BS antibodies isolated via flow sorting were reflective of the B cell receptor repertoires identified via the 10X genomics assays (**Figure 6D**), including predominant VH1-2 and VH1-46 gene usages with variable HCDR3 AA length and mutation frequencies (**Figure 7C, D, and E**). Interestingly, CH505TF gp120-reactive antibody Ab027646 utilizing VH3-15, not previously reported for usage by CD4BS antibodies, bound the monomeric CH505TF gp120 core protein^31^ lacking the V1V2 and V3 loops of Envs and is preferentially bound by CD4bs antibodies (**Figure 7F**) that are preferentially bound by CD4BS antibodies. Ab027646 showed a similar binding profile to WT and mutant CH505 core proteins as non-bnAb (but still neutralizing) F105 (**Figure 7F**). Unlike CH103-like antibodies, Ab027646 bound a core with the CD4bs disrupted (CH505TF gp120_STG) revealing an F105-like binding phenotype. Negative stain electron microscopy (NSEM) of Ab027646 in complex with CH505TF SOSIP when overlaid with F105 bound to the same trimer revealed overlapping binding footprints for both antibodies (**Figure 7G**). Both Ab027646 and F105 neutralized only vaccine-strain tier 1 CH505 w4.3, but failed to neutralize tier 2 CH505TF (in contrast to CH103 bnAb) (**Figure 7H**). Overall, these results confirmed vaccination with monomeric CH505TF gp120 induced CD4BS-like antibodies that were only capable of neutralizing Tier-1 strains.

## Discussion

The results from HVTN 115 demonstrate clearly that vaccination, with even the most promising low affinity gp120 monomers identified via extremely detailed natural infection studies of HIV Env sequences that lead to bNAbs, is insufficient to prime CD4bs bNAb lineages in adults. The vaccine was generally safe and well tolerated, including with repeated doses. It did not, however, convincingly expand either the CH103 or the CH235 lineages that were expanded during natural infection with exposure to the same antigen via vaccination. Additionally, the vaccine induced robust vaccine-specific B cell responses, vaccine-specific T cell responses, and non-neutralizing binding antibodies, suggesting that the lack of expansion of CH103 or CH235 lineages was not due to poor overall immunogenicity, and that GLA-SE is a reasonably potent adjuvant in the context of Env vaccination. From these data, we propose that further investigations in adults using recombinant Env should use different approaches other than low-affinity monomers.

Priming with CH505TF gp120 followed by boosting in sequential (T5) or additive (T6) regimens of CH505 gp120s from the stepwise development of the CH103 lineage induced only limited Tier 1A autologous neutralizing antibodies, and no substantial breadth or indication of bnAb precursor development. Our examination of the CH505TF gp120-induced BCR repertoire provided mechanistic insight into the developmental failure to recapitulate the CH103 lineage with this approach. Although B cells utilizing heavy chain variable regions associated with canonical CD4bs bnAbs (e.g. VH1-2, VH1-46, and VH4-59) were induced, the resulting CD4bs-specific mAbs they encoded for did not have the features consistent with VRC01 or CH103 bnAbs. Instead, the phenotype of the induced B cells was consistent with non-bnAb CD4bs antibodies such as F105 that favor binding of open-trimer conformations, and limited binding to more native-like closed trimer conformations^32^. Our data indicate that monomeric CH505TF gp120 was insufficient to induce detectable bnAb precursors due to inherent differences with the CD4bs presented on more native-like Env trimers. The repertoire of CD4bs antibodies elicited by CH505 TF gp120 in humans provides the opportunity to characterize these antibodies and use them in immunogen design to identify Envs that will disfavor the induction of off-target, non-neutralizing CD4bs antibodies. The concept would be to eliminate or obscure these epitopes in subsequent iterations.

Despite the lack of bnAb lineage priming, HVTN 115 provides important lessons on how to conduct clinical testing of regimens intended to elicit bnAbs, and has informed the next generation of HIV vaccine trials termed “discovery medicine^33^.” This study underscores the need to incorporate into clinical trial design clear bnAb lineage development targets and objective measures, such as BCR repertoire analysis coupled with structural analysis. Even though B cells with IgH genes identical to known CD4bs bnAb lineages were clearly enriched in vaccinated participants (VH1-2, and VH1-46, for VRC01 and CH235, respectively), these full sequences of isolated monoclonal antibodies suggested that they could never start this lineage development or develop functional properties required for broad neutralization. Additionally, monoclonal antibodies isolated from the CD4bs fraction of vaccine-reactive B cells had functional qualities more in line with Tier 1-neutralizing antibodies (e.g. would only be capable of the open trimer configuration typical of lab-adapted strains). In subsequent trials, the main objective should be to determine the effectiveness of a priming regimen in inducing sufficient bnAb precursors. This understanding will inform whether follow-up boosting strategies are warranted to further advance mature bnAb development. Additionally, data from Part A with repeated homologous administrations of the transmitted founder suggest that even repeated administrations with a monomeric gp120 cannot overcome an initial lack of bnAb priming in adults.

Since the inception of HVTN 115, and consistent with our findings of inadequate induction of bnAb precursors with monomeric gp120, there is a realization that effective bnAb lineage induction will require a different approach than vaccination with unmodified gp120 monomers. Subsequently, their design and production principles have substantially advanced and now a multitude of next-generation native-like trimer immunogens have entered clinical trials, including a high affinity version CH505TF (HVTN 300, NCT04915768). Additional iterations on this series of immunogens are being tested, including a membrane-bound construct (CH505M5 N197D) in HVTN 312. The B cell repertoire and transcriptome analyses of CH505TF gp120 provides a benchmark for comparison with ongoing trials designed to elicit CD4bs bnAb precursor antibodies.

Importantly, the establishment of a reasonable safety profile and dosing immunogenicity for CH505TF gp120 adjuvanted with 10 mcg of GLA-SE in Part A enabled its evaluation in HIV-exposed infants in South Africa in HVTN 135 (clinicaltrials.gov #NCT04607408), with lower doses of adjuvant^34^, to assess the suggested lower threshold for bnAb development that may be present in young children^35,36^. These results will be reported in a separate publication.

Overall, by meticulously assessing the potential of monomeric gp120 to induce the necessary precursors for CD4bs bnAb lineage development, HVTN 115 served as an important and necessary step in the clinical developmental pathway toward bnAb-inducing vaccines.

## Data Availability

De-identified participant-level and aggregate-level data will be made publicly available.

## Data sharing

De-identified participant-level and aggregate-level data will be made publicly available via the Atlas Science Portal at https://atlas.scharp.org/project/HVTN%20Public%20Data/begin.view.

## Acknowledgements

The authors would like to thank all participants of the trial, as well as all HVTN 115 study team members.

