## Supplemental Material for "A phase 1 randomized controlled trial to evaluate the safety and immunogenicity of a HIV monomeric gp120 protein B-cell lineage targeting HIV vaccine in healthy adults"

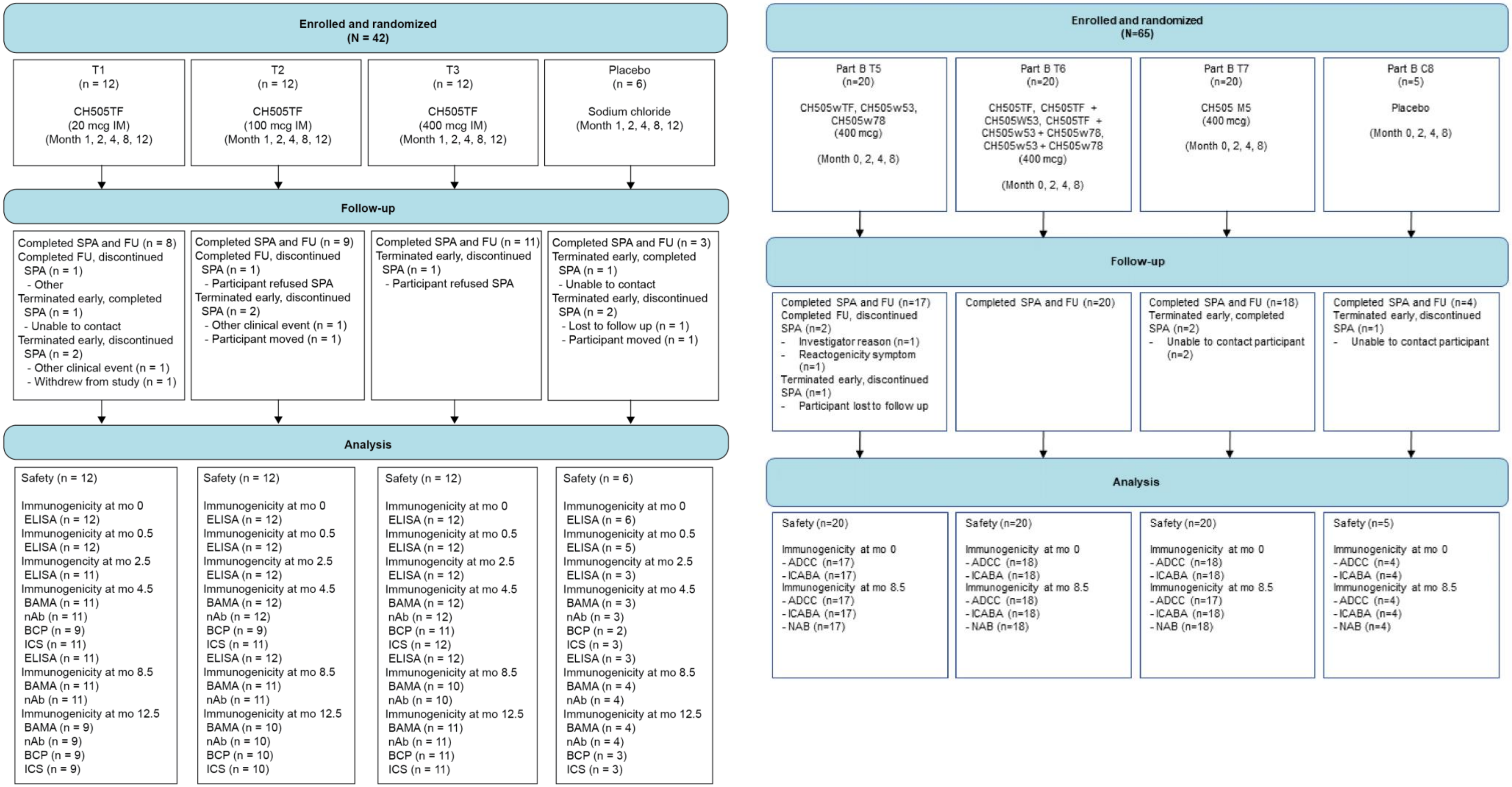

Supplemental Figure 1. CONSORT diagram of HVTN 115 Part A and B.

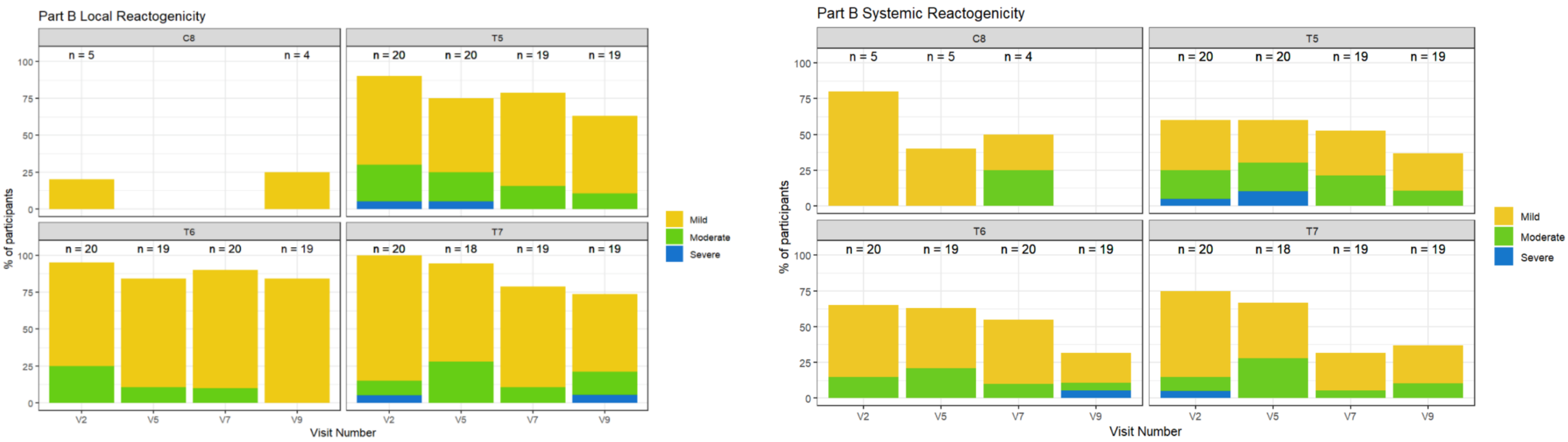

Supplemental Figure 2. Local and Systemic Reactogenicity for HVTN 115 Part B.

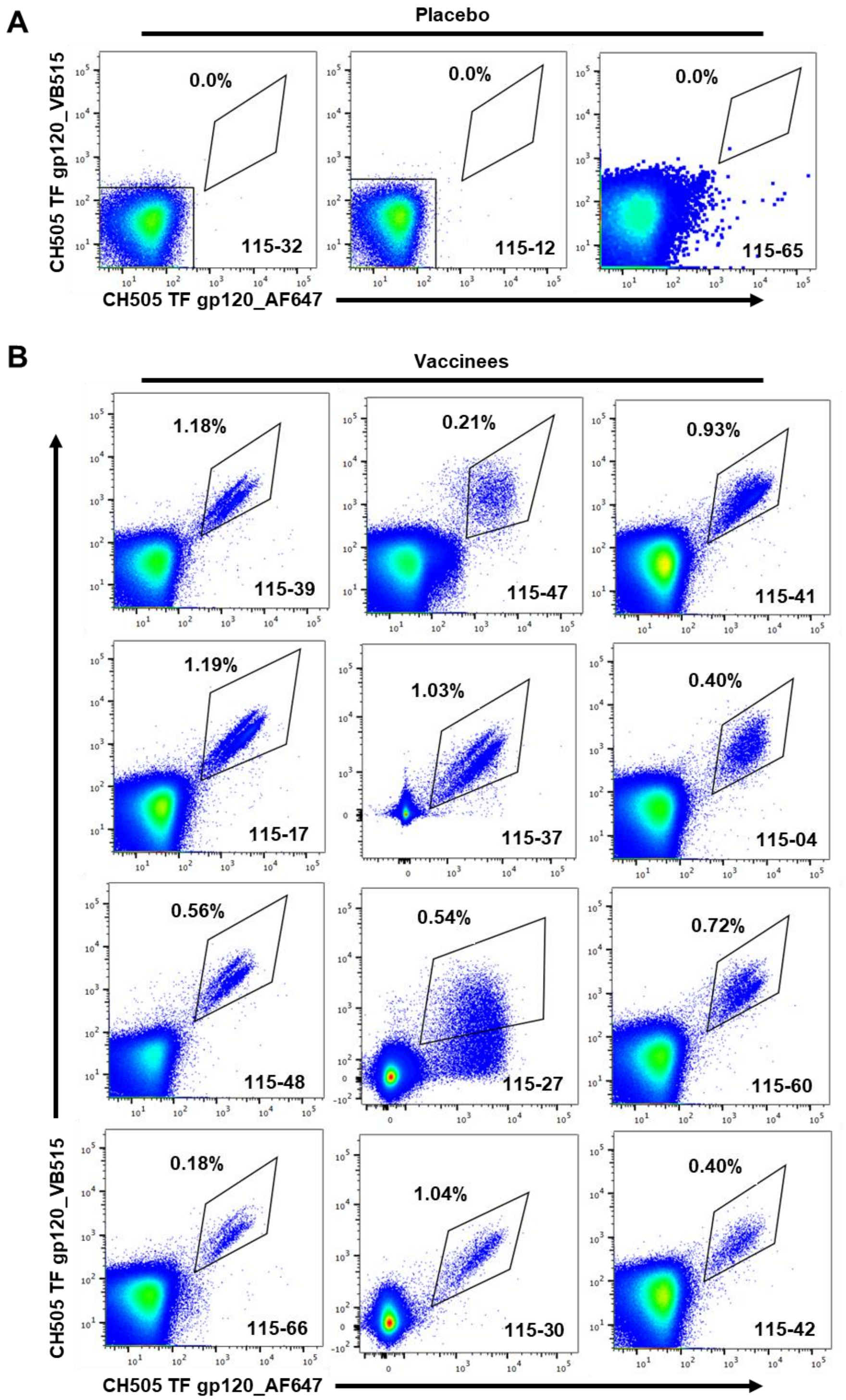

Supplemental Figure 3. A) Representative flow cytometry gating for sorting of CH505 TF gp120+ B cells from vaccinees post-fifth immunization (N=12) and antigen-negative B cells from placebo recipients (N=3).

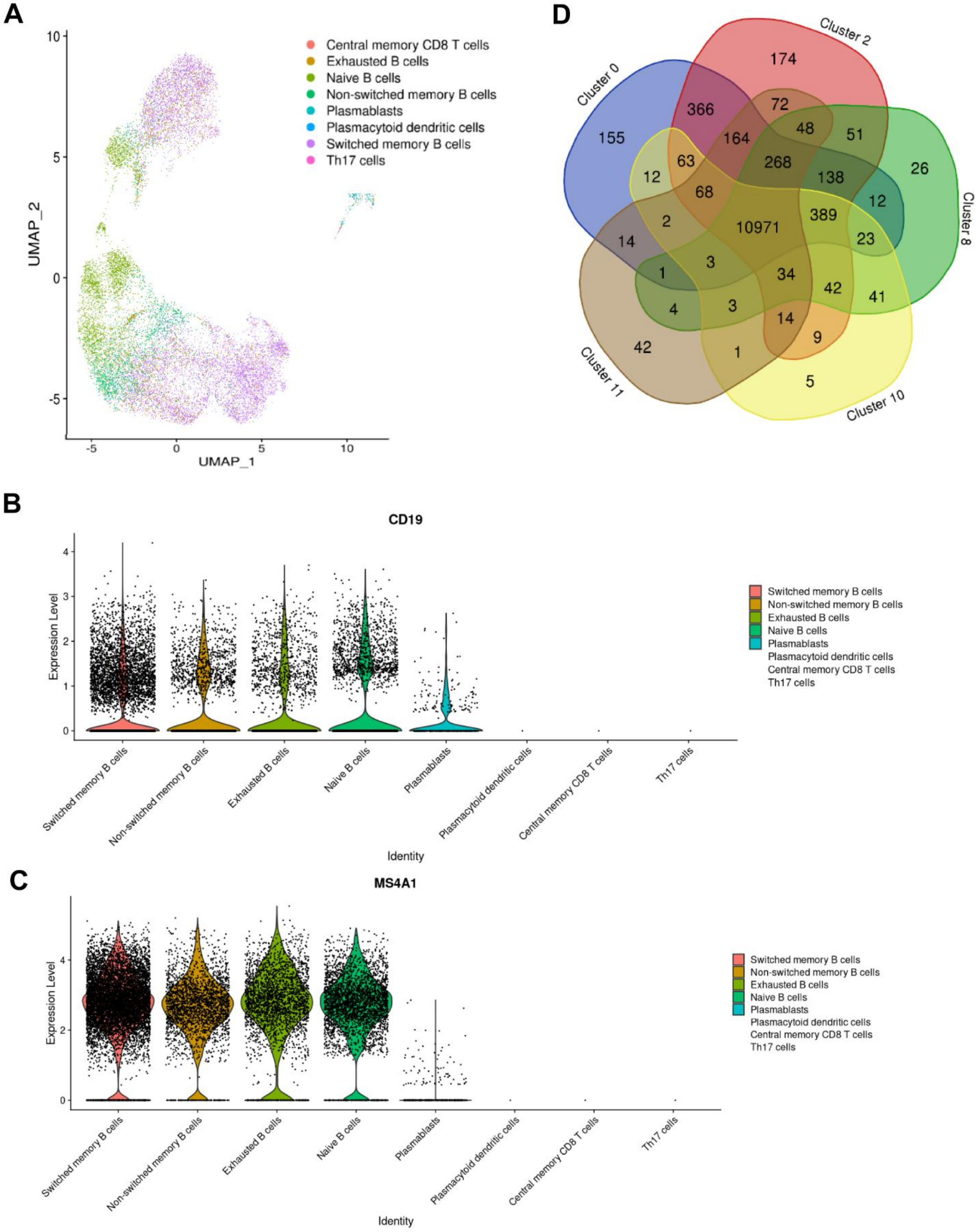

A
Central memory CD8 T cells
Exhausted B cells
Naive B cells
Non-switched memory B cells
Plasmablasts
Plasmacytoid dendritic cells
Switched memory B cells
Th17 cells
UMAP_1
UMAP_2
D
Cluster 0
Cluster 2
Cluster 8
Cluster 10
Cluster 11
B
CD19
Expression Level
Identity
C
MS4A1
Expression Level
Identity

Supplemental Figure 4. **A)** UMAP detailing inferred phenotypes associated gene expression profiles from antigen-reactive B cells. Expression of canonical B cell markers CD19 **(B)** and CD20 or MS4A1 **(C)** in each cluster of cells. The scarce number of cells inferred as non-B cells did not express CD19 or CD20, suggestive of cells that may have carried over during the sort of antigen-reactive B cells or artefact of singleR cluster inference. **D)** Venn diagram displayed the number of shared genes among the five transcriptionally unique clusters 0, 2, 8, 10, and 11 among the vaccine recipients. Numbers indicate the gene counts per cluster; overlap indicated common genes between clusters.

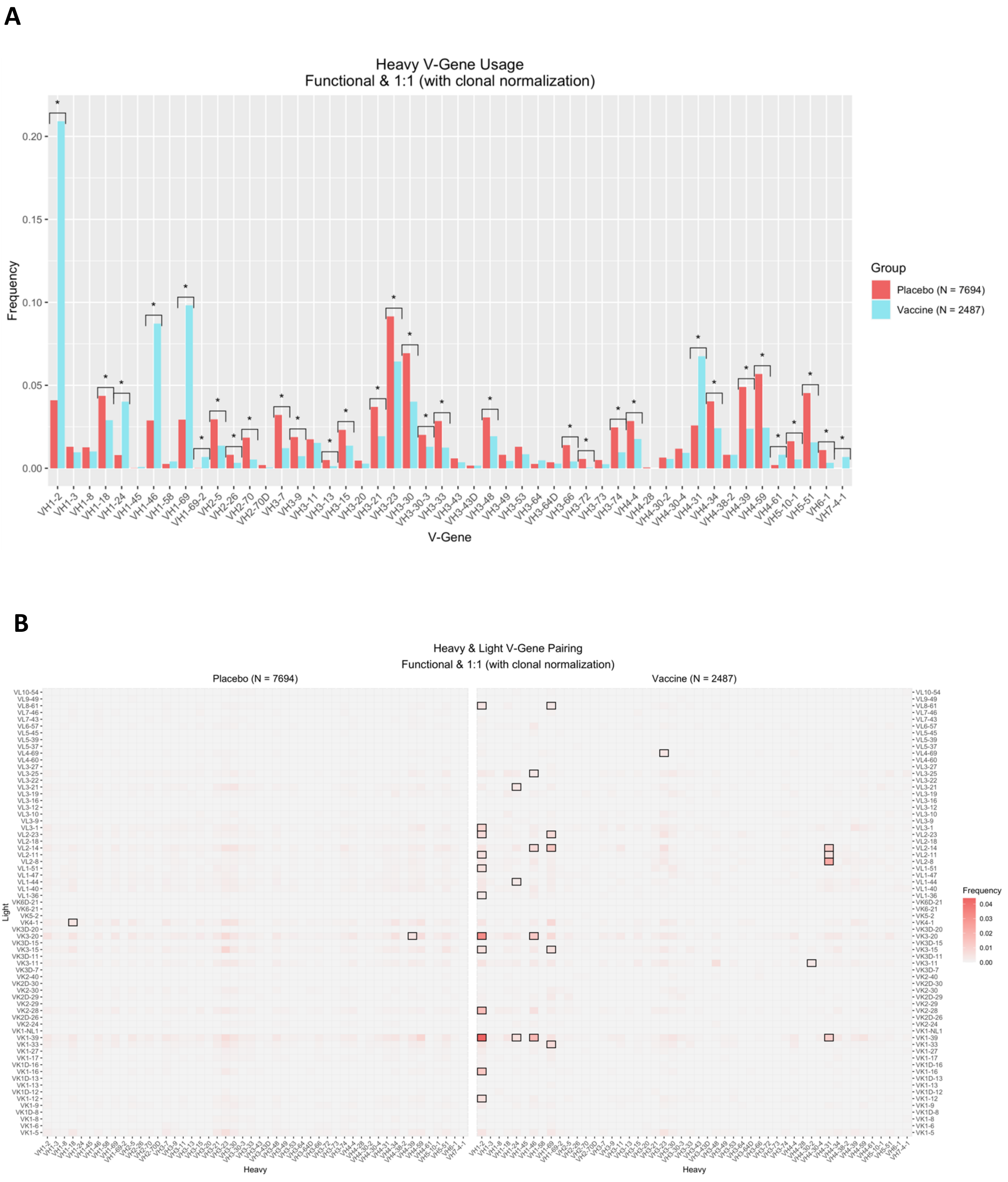
A
Heavy V-Gene Usage
Functional & 1:1 (with clonal normalization)
Frequency
0.20
0.15
0.10
0.05
0.00
V-Gene
VH1-2 VH1-3 VH1-8 VH1-18 VH1-24 VH1-45 VH1-46 VH1-58 VH1-69 VH1-69-2 VH2-5 VH2-26 VH2-70 VH2-70D VH3-7 VH3-9 VH3-11 VH3-13 VH3-15 VH3-20 VH3-21 VH3-23 VH3-30 VH3-30-3 VH3-33 VH3-43 VH3-43D VH3-48 VH3-49 VH3-53 VH3-64 VH3-64D VH3-66 VH3-72 VH3-73 VH3-74 VH4-4 VH4-28 VH4-30-2 VH4-30-4 VH4-31 VH4-34 VH4-38-2 VH4-39 VH4-59 VH4-61 VH5-10-1 VH5-51 VH6-1 VH7-4-1
Group
Placebo (N = 7694)
Vaccine (N = 2487)
B
Heavy & Light V-Gene Pairing
Functional & 1:1 (with clonal normalization)
Placebo (N = 7694)
Vaccine (N = 2487)
Light
Heavy
Frequency
0.04
0.03
0.02
0.01
0.00

C

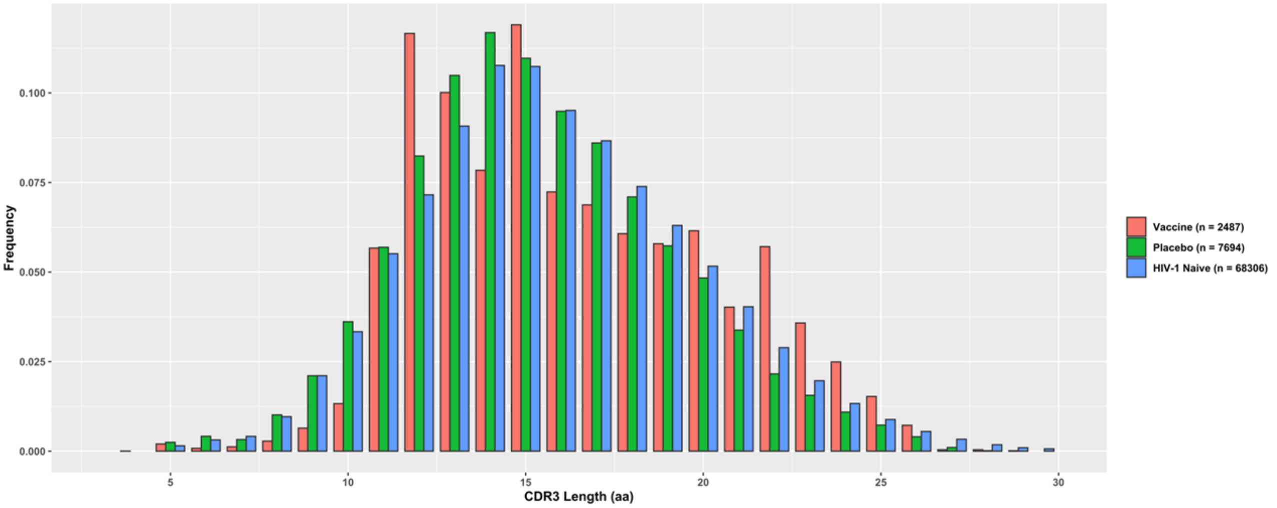

| Fisher's Exact Test | | |
|---|---|---|
| **Recipient** | **P-value** | **Adjusted p-value (FDR)** |
| ≥18 | 2.27e-17 | 2.59e-17 |
| ≥20 | 2.90e-29 | 8.69e-29 |
| ≥23 | 2.59e-17 | 2.59e-17 |

| Recipient | CDRH3 ≤ 18 AA | CDRH3 >18 AA |
|---|---|---|
| Placebo | 6154 | 1540 |
| Vaccine | 1739 | 748 |

| Wilcoxon | |
|---|---|
| **P-value** | 7.25e-17 |

D

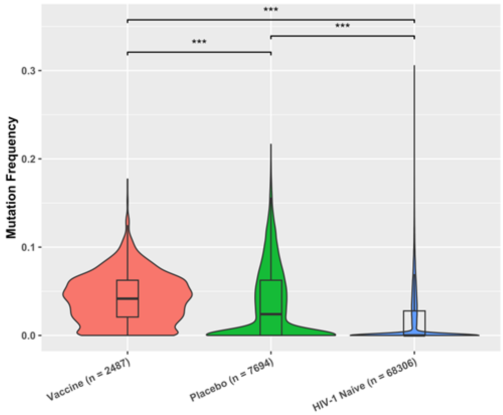

E

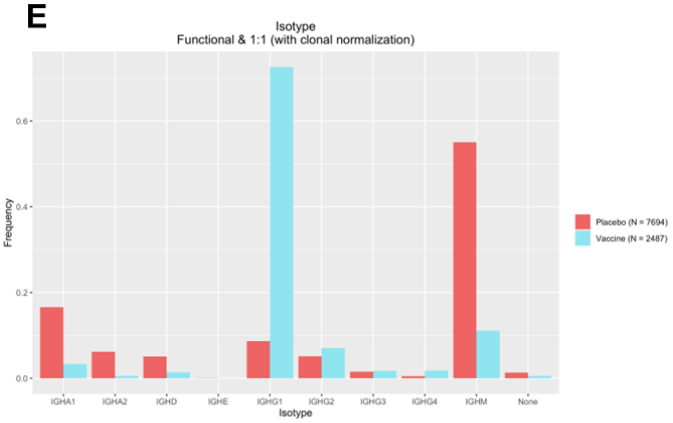

Supplemental Figure 5. A) Frequency of VH gene usage in antigen-reactive B cells in vaccinees (N=12) and placebo (N=3) individuals. B) Heat map of preferential heavy and light chain V-gene pairings from isolated vaccinee (N=2487) and placebo (N=7694) B cell receptors. C) HCDR3 length of isolated vaccinee (N=2487) and placebo (N=7694) B cell receptors. D) Heavy chain mutation frequency of isolated vaccinee (N=2487), placebo (N=7694), and HIV-1 naïve (68306) B cell receptors. E) Frequency of isotype usage from isolated vaccinee (N=2487) and placebo (N=7694) B cell receptors. Pv = *>0.05, **>0.01, ***>0.001, ****>0.0001.

**A**

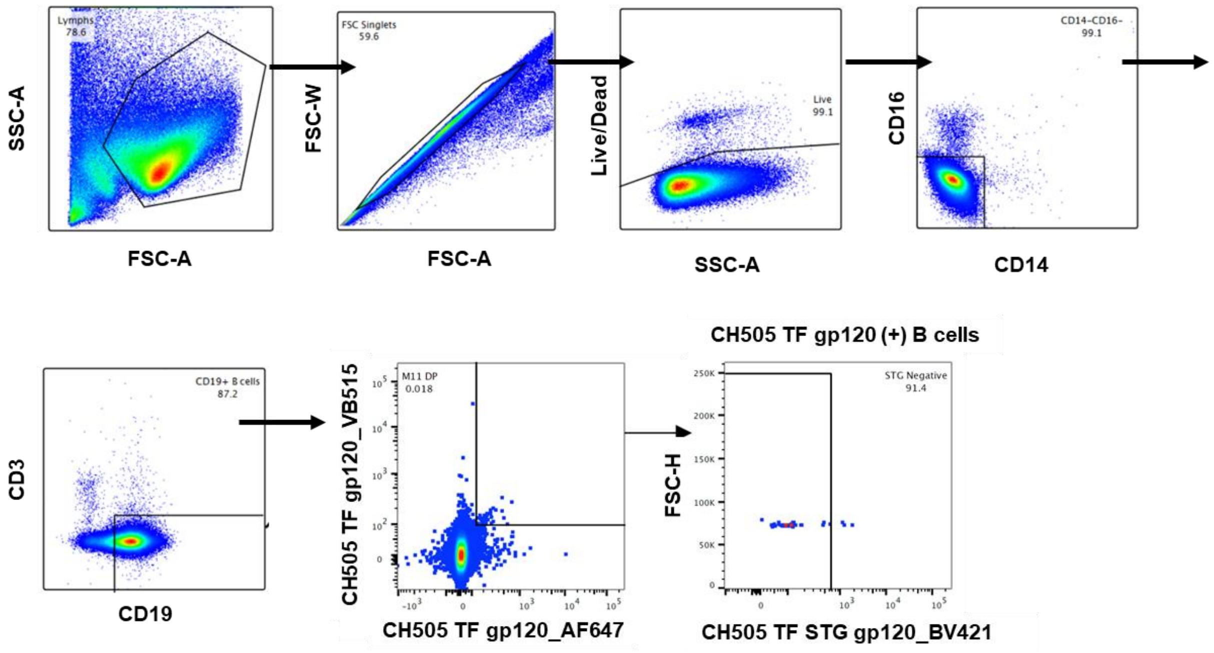

**B**

| | Total B cells | | | Memory B cells | | |
|---|---|---|---|---|---|---|
| Pub-ID | Cell count | CH505 SOSIP DP count | STG- count | Cell count | CH505 SOSIP DP count | STG- count |
| 115-27 | 1,229,961 | 19 | 17 | 507,831 | 11 | 10 |
| 115-42 | 275,482 | 0 | 0 | 36,935 | 0 | 0 |
| 115-48 | 479,018 | 7 | 4 | 126,165 | 3 | 1 |
| 115-66 | 112,575 | 3 | 2 | 21,470 | 1 | 0 |
| 115-43 | 267,376 | 8 | 6 | 67,093 | 3 | 1 |
| 115-35 | 205,557 | 6 | 4 | 49,803 | 1 | 1 |
| 115-39 | 49,267 | 1 | 0 | 19,134 | 0 | 0 |
| 115-37 | 743,668 | 19 | 14 | 145,863 | 14 | 11 |
| 115-04 | 395,747 | 9 | 8 | 62,918 | 5 | 5 |
| 115-47 | 422,005 | 10 | 10 | 58,352 | 4 | 4 |
| 115-17 | 843,519 | 27 | 27 | 229,192 | 17 | 17 |
| 115-60 | 192,367 | 35 | 32 | 60,105 | 27 | 24 |
| 115-30 | 276,686 | 9 | 8 | 54,052 | 5 | 5 |
| 115-41 | 375,166 | 411 | 410 | 109,122 | 319 | 318 |

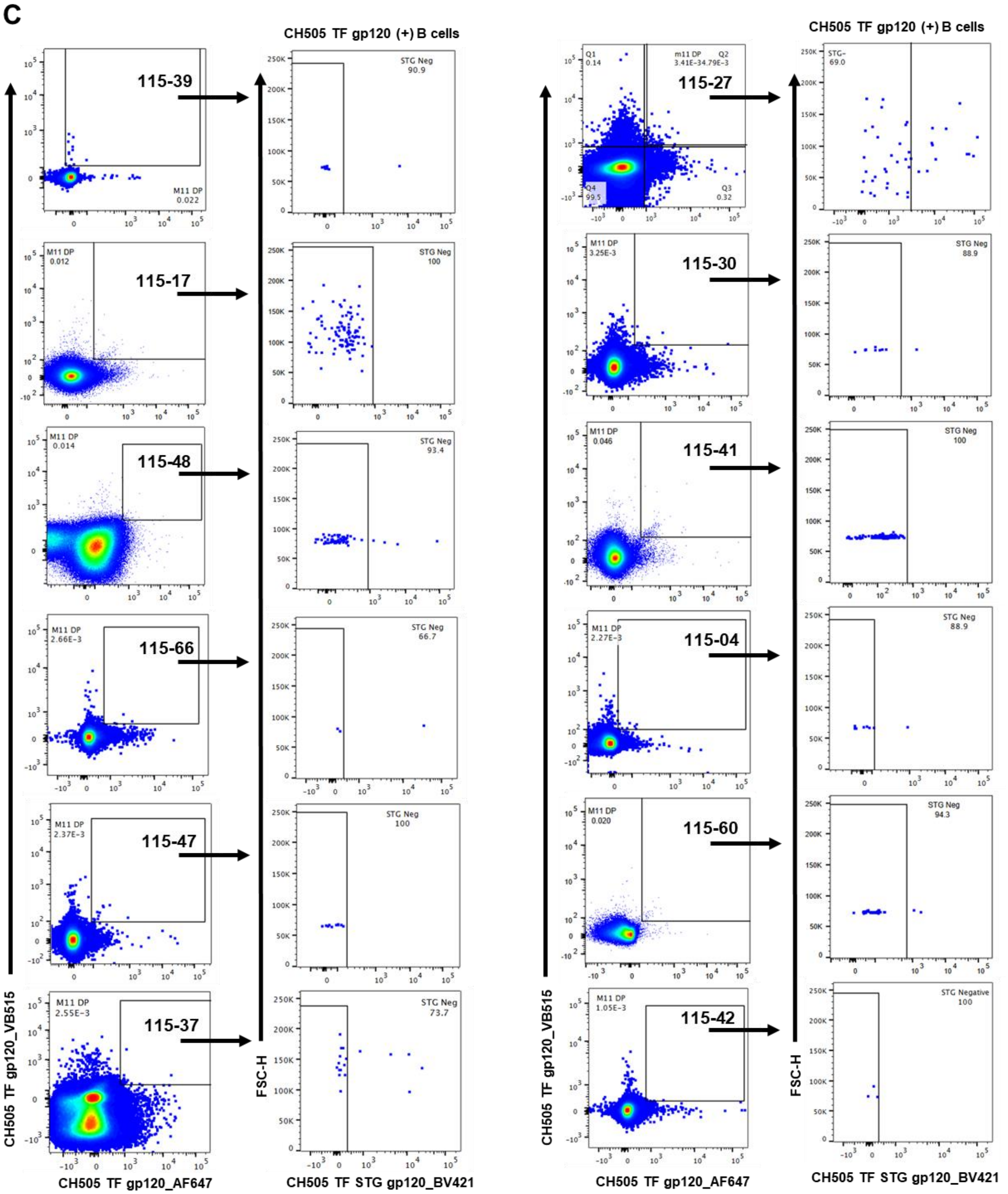

Supplemental Figure 6. A) Representative flow cytometry gating strategy for sorting of antigen-reactive CH505 TF gp120+ CH505 TF STG- B cells for single cell PCR from vaccinees post-third immunization. B) Numerical analysis from antigen-reactive CH505 TF gp120+ CH505 TF STG- B cell sorts (N=12). C) Representative flow cytometry gating for sorting of antigen-reactive CH505 TF gp120+ CH505 TF STG- B cells for single cell PCR from vaccinees post-third immunization (N=12).

| Supplemental Table 1. Serum IgG tested antigens | |
|---|---|
| **Full antigen name** | **Short name** |
| 1086C D7gp120.avi/293F | gp120 1086C |
| CH0505.w53.e16.D8gp120/293F | CH505.w53 |
| CH0505.w78.env33.D8gp120/293F | CH505.w78 |
| CH0505 CON D7gp120 D368R avi/293F/Mon | CH505 CON gp120 D368R |
| CH0505 CON D7gp120 avi/293F/Mon | CH505TF gp120 |
| CH505.M5D8gp120/293F | CH505.M5 gp120 |
| CH505TF D7gp120d371 avi/293F/Mon | CH505TF gp120 d371 |
| CH505TFD8N156KN160K avi/293i Mon | CH505TF gp120 N156KN160K |
| CH505TF D8gp120/293F | CH505TF gp120 |
| Con 6 gp120/B | gp120 B Con 6 |
| Con S gp140 CFI | gp140 Con S |
| gp41 | gp41 |
| gp70 CH505TF V1V2 N156KN160K avi/293F | CH505TF V1V2 N156KN160K |
| gp70 CH505TF V1V2 avi/293F | CH505TF V1V2 |
| CH505TF.6R.SOSIP.664.v4.1 avi.2Bio/293F | CH505TF SOSIP |

Supplemental Table 2. 17-color ICS staining panel.

| Antibody | Manufacturer | Catalog Number |
|---|---|---|
| AViD* (Viability marker) | Invitrogen | L34957 |
| CD3 BUV737 | BD Biosciences | 564307 |
| CD4 BUV395 | BD Biosciences | 563550 |
| CD8 BV650 | BD Biosciences | 563821 |
| CD14 BV510* | BioLegend | 301842 |
| CD56 BV570 | BioLegend | 318330 |
| CXCR5 PE-Dazzle594 | BioLegend | 356928 |
| PD-1 (CD279) BV605 | BioLegend | 329924 |
| ICOS (CD278) BV711 | BD Biosciences | 563833 |
| CD45RA APC H7 | BD Biosciences | 560674 |
| CCR7 BV786 | BioLegend | 353229 |
| IFNγ V450 | Becton Dickinson | 560371 |
| TNFα FITC | eBioscience | 11-7349-82 |
| IL2 PE | BD Biosciences | 559334 |
| IL4 PerCP-Cy5.5 | BioLegend | 500822 |
| IL17a PE-Cy7 | BioLegend | 512315 |
| CD40L APC | BD Biosciences | 555702 |
| Granzyme B Alx700 | BD Biosciences | 560213 |

Supplemental Table 3. Tally of B cells studied from HVTN 115 Part A participants via single cell immune profiling assay. Percent functional 1:1 B cells is the frequency of total B cells studied, and percent clonal normalization is a frequency of total B cells studied. B cells were studied at post 5th immunization as the final immunization in all participants, except 115-42 where we studied B cells post 3rd immunization.

| Group | Pub ID | Number of Cells | Number of Cells with Productive V-J Spanning Pair | Functional & 1:1 B Cells | Number of B Cells After Clonal Normalization |
|---|---|---|---|---|---|
| Vaccine recipients | 115-27 | 2305 | 1398 (60.7%) | 1090 (47.3%) | 394 (17.1%) |
| | 115-60 | 966 | 779 (80.6%) | 721 (74.6%) | 350 (36.2%) |
| | 115-66 | 216 | 158 (73.1%) | 138 (63.9%) | 98 (45.4%) |
| | 115-30 | 205 | 165 (80.5%) | 137 (66.8%) | 97 (47.3%) |
| | 115-48 | 271 | 141 (52.0%) | 128 (47.2%) | 88 (32.5%) |
| | 115-41 | 1021 | 894 (87.6%) | 768 (75.2%) | 340 (33.3%) |
| | 115-04 | 40 | 21 (52.5%) | 20 (50.0%) | 15 (37.5%) |
| | 115-39 | 532 | 215 (40.4%) | 187 (35.2%) | 130 (24.4%) |
| | 115-17 | 1696 | 940 (55.4%) | 824 (48.6%) | 253 (14.9%) |
| | 115-37 | 2027 | 1396 (68.9%) | 1146 (56.5%) | 459 (22.6%) |
| | 115-47 | 338 | 279 (82.5%) | 212 (62.7%) | 167 (49.4%) |
| | 115-42 | 151 | 122 (80.8%) | 107 (70.8%) | 96 (63.6%) |
| | Group Total | 9768 | 6508 (66.6%) | 5478 (56.1%) | 2487 (25.5%) |
| Placebo recipients | 115-12 | 3467 | 2576 (74.3%) | 2231 (64.3%) | 1816 (52.4%) |
| | 115-65 | 3199 | 2715 (84.9%) | 2286 (71.5%) | 2065 (64.6%) |
| | 115-32 | 7426 | 6105 (82.2%) | 4743 (63.9%) | 3813 (51.3%) |
| | Group Total | 14092 | 11396 (80.9%) | 9260 (65.7%) | 7694 (55.0%) |
| All | Total | 23860 | 17904 (75.0%) | 14738 (61.8%) | 10181 (42.7%) |

Supplemental Tables 4 and 5. **(see separate Excel files)**
